# A Flexible Statistical Framework for Estimating Excess Mortality

**DOI:** 10.1101/2020.06.06.20120857

**Authors:** Rolando J. Acosta, Rafael A. Irizarry

## Abstract

Quantifying the impact of natural disasters or epidemics is critical for guiding policy decisions and interventions. When the effects of an event are long-lasting and difficult to detect in the short term, the accumulated effects can be devastating. Mortality is one of the most reliably measured health outcomes, partly due to its unambiguous definition. As a result, excess mortality estimates are an increasingly effective approach for quantifying the effect of an event. However, the fact that indirect effects are often characterized by small, but enduring, increases in mortality rates present a statistical challenge. This is compounded by sources of variability introduced by demographic changes, secular trends, seasonal and day of the week effects, and natural variation. Here we present a model that accounts for these sources of variability and characterizes concerning increases in mortality rates with smooth functions of time that provide statistical power. The model permits discontinuities in the smooth functions to model sudden increases due to direct effects. We implement a flexible estimation approach that permits both surveillance of concerning increases in mortality rates and careful characterization of the effect of a past event. We demonstrate our tools’ utility by estimating excess mortality after hurricanes in the United States and Puerto Rico. We use Hurricane Maria as a case study to show appealing properties that are unique to our method compared to current approaches. Finally, we show the flexibility of our approach by detecting and quantifying the 2014 Chikungunya outbreak in Puerto Rico and the COVID-19 pandemic in the United States. We make our tools available through the `excessmort` R package available from https://cran.r-project.org/web/packages/excessmort/.

## Introduction

Accurate and timely estimation of all-cause mortality rates after a natural disaster or infectious disease outbreak is paramount as it serves as a way to quantify health effects, aid in policy making, and resource allocation. The US Center for Disease Control and Prevention (CDC) defines a directly related disaster death as one that is attributable to the forces of the disaster or by consequences of these forces. A death indirectly related to a disaster occurs when unsafe or unhealthy conditions present during any phase of the disaster contribute to it[1]. In the case of epidemics, lack of comprehensive testing or reporting can lead to challenges in measuring direct effects, while indirect effects can arise due to, for example, increased stress levels or reduced access to health services.

Excess mortality, defined as the subtraction of the expected number of deaths from the observed counts in a period of interest[2], is frequently estimated using historical data and a model that accounts for seasonal and secular trends, as well as demographic changes. For example, to estimate expected counts in Puerto Rico and assess the impact of Hurricane Maria, various groups have used mortality data to estimate excess deaths[3, 4, 5, 6]. More recently, methods for estimating excess mortality have been used to assess the impact of the COVID-19 pandemic[7, 8, 9]. The CDC uses an adaption of an outbreak detection method known as the Farrington algorithm to estimate excess deaths[10, 11, 12].

The effects of natural disasters and epidemics typically last longer than a week and can also change within a week. In the case of natural disasters, we might see a sharp increase in the death rate on the day of the event, followed by a smooth decline back to normal levels lasting several weeks. When indirect effects are severe, this decline will be slow. For epidemics, the patterns might be characterized by a short period of exponential growth followed by a plateau and a decline back to normal. In both cases, we expect the decline to be a smooth function of time, often of hard to detect magnitude, and incorporating this into a model can provide statistical power. We sought to develop a flexible approach that leverages the availability of daily data to effectively estimate these smooth trends and sharp increases. Specifically, we extended and improved previously published methods based on Poisson regression [11, 12, 13, 14] by modeling the event effect as a smooth function of time. Because natural variability introduces more variance than predicted by a Poisson model, and because daily data exhibits correlation in time, we proposed a mixed effects model that includes an auto-regressive process.

We use a simulation study to demonstrate that our approach provides accurate and precise estimates. To demonstrate the utility of our approach we searched for periods of excess mortality during the last 35 years in Puerto Rico, detecting large effects during the 2014 Chikungunya outbreak and after hurricanes Georges and Maria. We demonstrate the advantages of our approach over current ones by examining the different estimates obtained for Hurricane Maria and Georges. We then compare the effect of these hurricanes in Puerto Rico to the effects of three other major hurricanes in the United States. Finally, we examined excess mortality during the COVID-19 pandemic in the United States. We make our method and wrangled datasets available through the `excessmort` R package. The code to reproduce the results presented here is available from GitHub: https://github.com/RJNunez/excess-mortality-paper

## Methods

### Statistical model

We modeled daily death counts with the following mixed model:

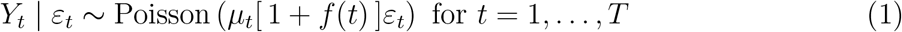

with *μ*_*t*_ the expected number of deaths at time *t* for a typical period, 100 *× f* (*t*) the percent increase at time *t* due to an unusual event, *ε*_*t*_ a time series of auto-correlated random variables representing natural variability, described in more detail in the eAppendix, and *T* the total number of observations.

The expected counts *μ*_*t*_ can be further decomposed into

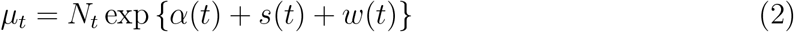

with *N*_*t*_ the population at time *t* that we treat as an offset, *α*(*t*) a slow-moving trend that accounts for secular changes such as the improved health outcomes we have observed during the last several years, *s*(*t*) a yearly periodic function representing a seasonal trend, and *w*(*t*) a day of the week effect. Note that factorizing the *N*_*t*_, rather than absorbing it into *α*(*t*), *s*(*t*), and *w*(*t*), does not change our approach to estimating or interpreting *f* (*t*), but permits us to interpret *α*(*t*) as a mortality rate and makes it comparable across groups of different population sizes. This is particularly useful in a jurisdiction like Puerto Rico where the population size has been decreasing substantially and with the working age population decreasing more than other age groups. In the eAppendix we provide details on how we obtain *N*_*t*_ for different jurisdictions.

We assume *α*(*t*) is smooth that is close to linear within 7 year periods. We further assume that the seasonal trend *s*(*t*) follows a harmonic model and we model the weekday-specific effects using seven indicator variables and seven constrained parameters. The details are included in the eAppendix.

We refer to *f* as the event effect and assume *f* (*t*) = 0 for typical periods not affected by natural disasters or outbreaks. When different from 0, we assume *f* (*t*) is smooth enough to be represented by a smoothing cubic spline with 12 knots per year. This provides enough flexibility to detect both natural disasters and outbreaks. If we know an event, such as a hurricane, occurred on a specific day, say *t*_0_, that could result in a sharp increase in death rate due to a direct effect, we permit a discontinuity at *t*_0_ to account for a sudden direct effect and fit a smoother spline, with 6 knots per year, to provide more power to detect subtle indirect effects. More details on these choices can be found in the Tuning parameters eAppendix section.

### Estimating event effects

Due to the flexibility in *f* (*t*) and the correlation structure of *ε*_*t*_, in practice, obtaining, for example, maximum likelihood estimates (MLE) for this model is not straightforward. To overcome this challenge, we implemented a three-step approach that works well in practice, as demonstrated by simulation and empirical validation described in the Results section. The general idea is to first estimate *μ*_*t*_ and the correlation structure of *ε*_*t*_ during periods with no events, referred to here as control periods, and then estimate the most interest component, *f* (*t*), assuming these are known. We use the Central Limit Theorem approximation to assume 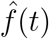 follows a normal distribution and compute standard error estimates 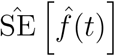 that include the variability introduced by the uncertainty in the estimate of the expected mortality rate 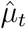. The details are described in the eAppendix.

### Detecting periods of concern

With the estimates 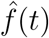 and 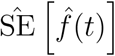 in place, we can construct a surveillance algorithm to detect periods of concern. We do this by grouping consecutive time points for which a percent increase of 0 is not in a 95% confidence interval for 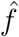. Specifically, we define concerning periods as [*t*_0_, *t*_1_] for which

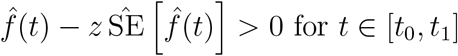

where we set *z* = 1.96, the 97.5 percentile of a standard normal distribution. We can increase specificity by increasing the degree of confidence to a percentage higher than 95% or by requiring an interval size of certain length before declaring a period concerning. The surveillance approach can either prioritize targeting short periods or long periods by using more or less knots, respectively. These can all be controlled in the R package through function arguments. For example, the `knots_per_year` in the function `excess_model` controls the smoothness of 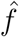. In the Results section we describe the power and false discovery rates for this procedure using simulation studies.

### Excess mortality estimate

Once we have identified a period of interest, either by surveillance or because we know an adverse event occurred, we can characterize the effect of this event by computing the cumulative excess death for the period. Note that we can conveniently represent excess deaths at time *t* as *μ*_*t*_*f* (*t*) and estimate these with 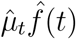. We can then estimate excess deaths for any time period [*t*_0_, *t*_1_] by just adding these up:

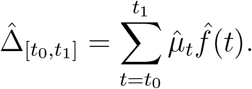

Note that we can use the formula for computing the sum of correlated random variables to estimate the standard error of 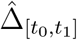. The details are included in the eAppendix.

### Mortality data

We obtained individual-level mortality records with no personal identifiers from the Department of Health of Puerto Rico’s Demographic Registry from January 1985 to August 2020. Using these, we computed daily death counts. Given Puerto Rico’s changing demographics[15], these counts were computed for six different age groups: 0 to 4, 5 to 19, 20 to 39, 40 to 59, 60 to 74, and 75 and older. We also obtained daily death counts from Florida, New Jersey, and Louisiana’s Vital Statistic systems from January 2015 to December 2018, January 2007 to December 2015, and January 2003 to December 2006, respectively. We further obtained weekly mortality counts from January 2017, to November 2020, made public on May 2020 by the CDC[10]. The CDC provides two types of mortality counts. First the counts reported by the states and second a weighted count intended to account for the lag in reporting[10]. Finally, we obtained COVID-19 mortality data for the US made public by the New York Times[16].

## Results

### Assessment via simulation study

To assess our procedure we conducted a Monte Carlo simulations. We designed simulation studies to mimic three scenarios 1) a natural disaster, 2) an infectious disease epidemic, and 3) a typical period with no events. The details of the simulations are in the eAppendix. We found that our method consistently estimates the true curve *f* (*t*) precisely under all three scenarios (eFigures 1A-C) and that our estimated standard error also estimates the true standard error precisely (eFigures 1D-F).

One of the advantages of our approach is that modeling the event effects as smooth functions greatly improves power over considering each time point individually. This is particularly powerful in scenarios in which low counts result in data with high coefficients of variation. To demonstrate this, and to determine how low counts-per-day rate our approach can handle, we repeated the above simulation but changing the average level of *α*(*t*) so that the daily rates were very low: 0.05, 0.10, 0.50, and 1.00 deaths per day. Our estimation procedure yielded precise and accurate estimates of the event effect when the rate was 0.10 or higher, while for 0.05 we started to see loss of accuracy (eFigures 2 & 3, eTable 1).

We also used these simulations to assess the sensitivity and specificity (false discovery rate) of our procedure for detecting excess mortality events. We examined several strategies for detecting periods of concern by varying the level of smoothness and the number of consecutive time points required to define the period as concerns as described in Section. Specifically, we considered three smoothing approaches, 1) smoothing with 6 knots per year, smoothing with 12 knots per year, and 3) a saturated model (no smoothing), and seven period length requirements: 1, 3, 5, 10, 30, and 60 days. We found that smoothing greatly improves sensitivity without much loss in specificity (eTables 2 & 3). As expected increasing the period length requirement increased specificity and reduced sensitivity. More details are included in the eAppendix.

Finally, to assess the susceptibility of our approach to different control periods used to estimate the expected rate *μ*_*t*_, we performed simulation and cross-validation studies. First, we evaluated the precision and accuracy of our estimate 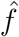 when estimating *μ*_*t*_ with 2, 4, 6, and 8 years of data. We found that performance was not affected (eFigures 4 & 5, eTable 4). We also ran a cross-validation using Puerto Rico data from 1999-2013 in Puerto Rico, fifteen consecutive years for which we do not expect to see a significant event. Specifically, we removed each year, one by one, estimated *μ*_*t*_ without that year and compared it to the estimate obtained when including that year for the estimation. We found almost no difference in the estimates (eFigure 6) demonstrating the robustness of our approach. More details on these simulation studies are included in the eAppendix.

### Comparison to the Farrington model

The current method implemented by the CDC is based on weekly totals:

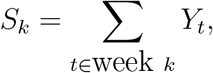

with *k* denoting week. They estimate the expected value and standard deviation for each *S*_*k*_ by applying the Farrington approach[11, 12] on historical data, what we call a control period. A threshold for anomalous mortality, *U*_*k*_, is defined as the upper bound of a one-sided 95% prediction interval:

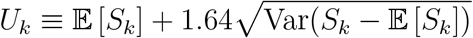

where 1.64 is the 95 percentile of a standard normal distribution. It follows that if *S*_*k*_ *> Û*_*k*_, then week *k* is denoted as having excess deaths. For each week, two estimates are provided: a conservative estimate, *S*_*k*_ *− Û*_*k*_, and an unbiased estimate, 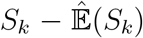, with 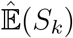 the estimated expected count at week *S*_*k*_.

To compare this method with our approach, we used Puerto Rico data that included the landfall of Hurricane Maria on the island. This category 4 hurricane interrupted the water supply, electricity, telecommunication networks, and access to medical care for several weeks [17, 18]. The consensus is that indirect effects of the storm were observed at least until December with well over 1,000 excess deaths[19]. However, the CDC approach only identifies four successive weeks with deaths above the threshold, which results in excess death estimates of 527 and 796 for the conservative and unbiased approaches, respectively (Figure 1A). We note that observed counts are above the expected value, 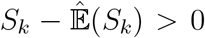, for six months after the storm, but only 19 of these are over the threshold, *S*_*k*_ *− Û*_*k*_ *>* 0. These data support a sustained indirect effect, yet the Farrington model lacks the power to detect the small and persistent increases in death rate introduced by such effects as one contiguous period. Applying the Farrington algorithm to daily data resulted in similar results (eFigure 7).

**Figure 1:**
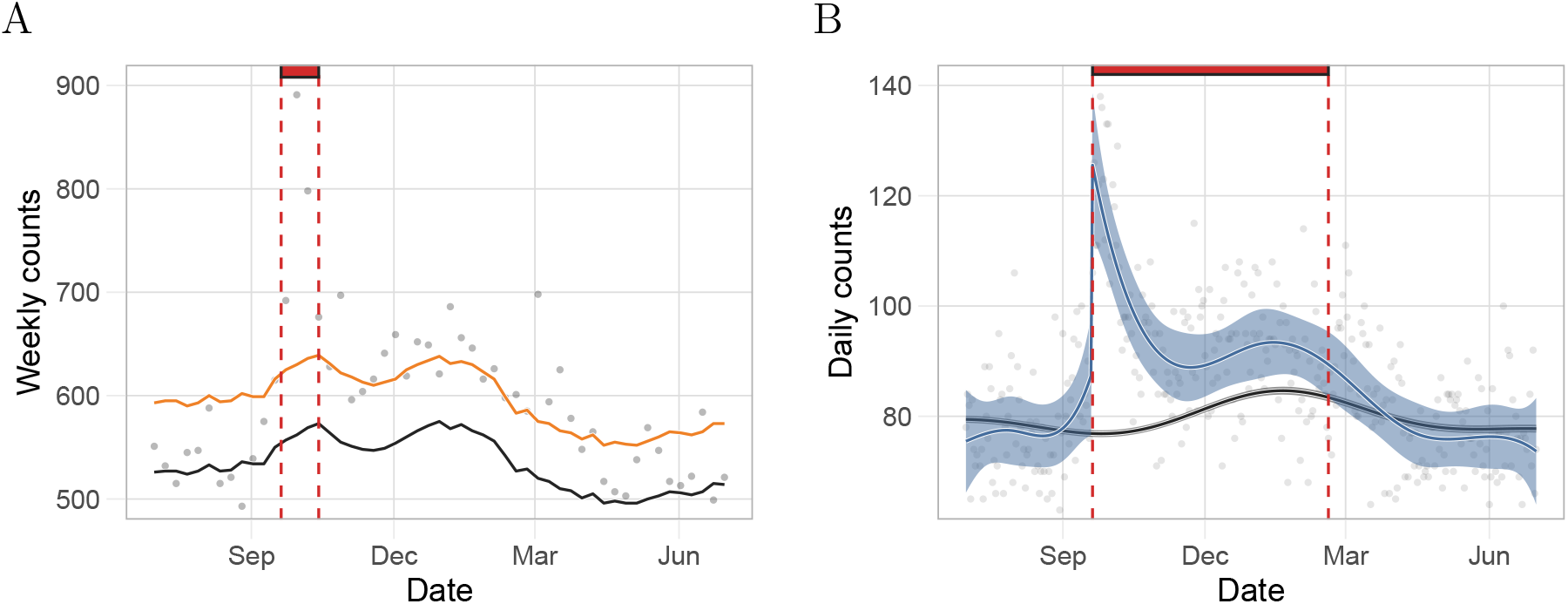
Comparison of our approach to the Farrington’s based on estimates for Puerto Rico from a period including Hurricane Maria. A) Gray points represent weekly deaths counts used by CDC. The black and the orange curves are the expected number of daily counts and the threshold for significant excess deaths, respectively, as defined by the Farrington algorithm. The red rectangle denotes the number of consecutive days with excess deaths since the landfall of Hurricane Maria as determined by the Farrington algorithm. B) Gray points represent daily death counts. The black curve is the estimated expected counts based on our method and the blue curve represents the event effect estimate, 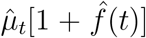. The black and blue ribbons are point-wise 95% confidence intervals for the expected counts and event effect, respectively. Finally, the red rectangle is as in A) but for our method.

We then fit our model to the same data, using the same estimates of population. Our approach was able to capture a sustained indirect effect (Figure 1B). Specifically, when we applied our approach to data from the same period, we found that a point-wise 95% confidence interval for 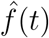 did not include zero for 151 consecutive days. This results in a substantially higher excess death mortality. Applying both of these approaches to data from Puerto Rico after Hurricane Georges in 1998 further illustrated the advantages of our approach (eFigures 8). In the next section we repeat this analysis but using the population size estimate described in Population size estimates section of the eAppendix and stratifying by age groups. We present an excess mortality estimate above 3,000.

### Quantifying indirect effects after hurricanes

As an example of the utility of our approach, we quantified and compared the direct and indirect effects of three hurricanes in Puerto Rico: Hurricane Maria in 2017, Hurricane Georges in 1998, and Hurricane Hugo in 1989. To assess if indirect effects are worse in Puerto Rico, we estimated and compared the impact of hurricanes in three other US jurisdictions: Hurricane Irma in Florida in 2017, Hurricane Sandy in New Jersey in 2012, and Hurricane Katrina in Louisiana in 2005 (Table 1, Figure 2 and eFigure 9). Since age-stratified data was available for Puerto Rico, we fit model (1) to each age group. Then, we aggregated the age group-specific effects to obtain the marginal effect of each hurricane (see eAppendix for details).

**Table 1:** Comparison of direct and indirect effects for six hurricanes. The first column shows the hurricane name and category. The second column shows the jurisdiction for which we obtained data. The third column is the date the hurricane made landfall. The fourth column is our estimate of the percent change in mortality rate the day after landfall including a 95% confidence interval. The fifth column is our estimate for the duration of indirect effects in days. The sixth column shows the excess mortality estimate and a 95% confidence interval for the period defined by column five. CI indicates confidence interval. ^*a*^The reported period for Katrina included 17 days with 0 in the 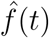 95% confidence interval.

| Hurricane<br>(Category) | Jurisdiction | Landfall<br>date | Percent<br>increase on<br>landfall (CI) | Indirect<br>effect<br>duration (days) | Excess<br>death<br>estimate (CI) |
| --- | --- | --- | --- | --- | --- |
| Hugo (3) | Puerto Rico | Sep. 18, 1989 | 16 (4 to 27) | 12 | 94 (24 to 163) |
| Georges (3) | Puerto Rico | Sep. 21, 1998 | 37 (25 to 49) | 90 | 1,300 (1,040 to 1,550) |
| María (5) | Puerto Rico | Sep. 20, 2017 | 74 (60 to 88) | 197 | 3,280 (2,890 to 3,670) |
| Katrina (5) | Louisiana | Aug. 29, 2005 | 718 (704 to 732) | 109<sup>a</sup> | 1,570 (1,300 to 1,830)<sup>a</sup> |
| Sandy (4) | New Jersey | Oct. 29, 2012 | 12 (3 to 22) | 12 | 195 (48 to 342) |
| Irma (5) | Florida | Sep. 10, 2017 | 6 (1 to 10) | 48 | 1,280 (790 to 1,760) |

**Figure 2:**
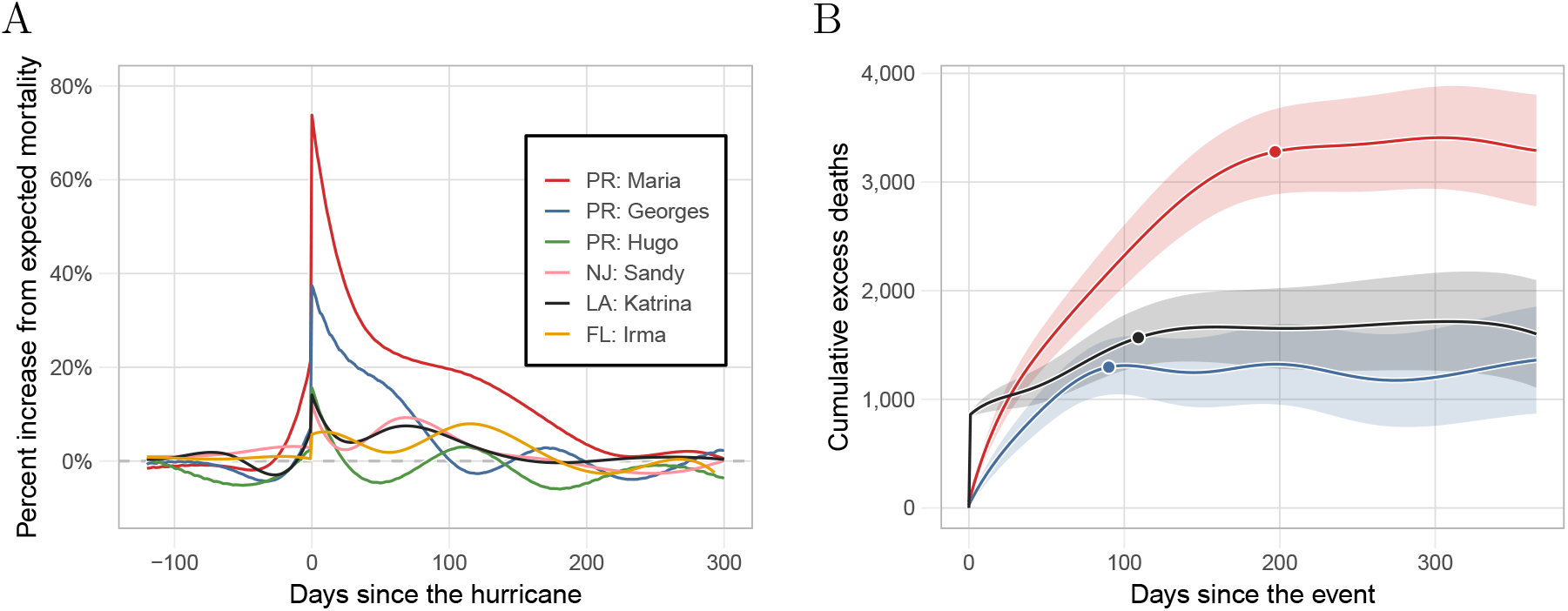
Estimated hurricane effects. A) Percent increase over expected mortality for the six hurricanes. B) Cumulative excess deaths for the 365 days after landfall for Hurricanes María, Georges, and Katrina, the three hurricanes with indirect effects lasting over two months. The data points correspond to the cumulative excess death estimate for the period of indirect effect presented in Table 1.

For Hurricane Maria in Puerto Rico, we found an increase in mortality of 74% (95% CI: 60% to 88%) on landfall and over 3,000 excess deaths in the subsequent months. These findings confirm previously reported results that the effects of hurricane Maria on Puerto Rico were unprecedented[3, 20]. For Georges, we found a similar yet less severe pattern to Maria. Specifically, we estimated an increase in mortality of 37% (95% CI: 25% to 49%) and over a thousand excess deaths in the three months after the storm. Conversely, the effects of Katrina on Louisiana were much more direct. On August 29, 2005, the day the levees broke, there were 834 deaths (data point not included in Figure 2A nor in eFigure 9E), which translates into a 718% (95% CI: 704% to 732%) increase in death rate. However, the increase in mortality rate for the ensuing months was substantially lower than in Puerto Rico after Maria and Georges (eFigure 9). The effects of Sandy on New Jersey and Irma on Florida were much less severe. For example, we estimated over 1,200 excess deaths in Florida after Irma which is on par with the excess death estimates associated with Hurricanes Irma and Hugo. However, note that the population of Florida is seven and five times larger than that of Puerto Rico and Louisiana, respectively. Furthermore, for both Florida and New Jersey, we see a second period of increase in mortality from December to March after each storm, respectively. These periods are consistent with the particularly bad Flu seasons of 2012 and 2018 (eFigure 9). We, therefore, do not include those periods as affected by the hurricane.

### Detecting and quantifying epidemics

As an example of how our approach can be used to detect and quantify the effects of epidemics or outbreaks, we fit our model to Puerto Rico mortality data from 1985 to 2020 and, apart from the hurricane seasons mentioned in the previous section, we detected an unusual increase in mortality rates from August 2014 to February 2015. This period coincides with the 2014-2015 Chikungunya outbreak[21, 22] (Figure 3A). The effects were particularly strong for individuals over 60 years and for the 0 to 4 years age group (eFigure 10). Cumulative excess mortality increased until February 2015, followed by a decrease in the ensuing months, consistent with a harvesting effect [23, 24]. A year after the start of the Chikungunya epidemic, on 1 August 2015, we observe a point estimate for excess mortality of 640 (95% CI: 140 to 1,140) (Figure 3B).

**Figure 3:**
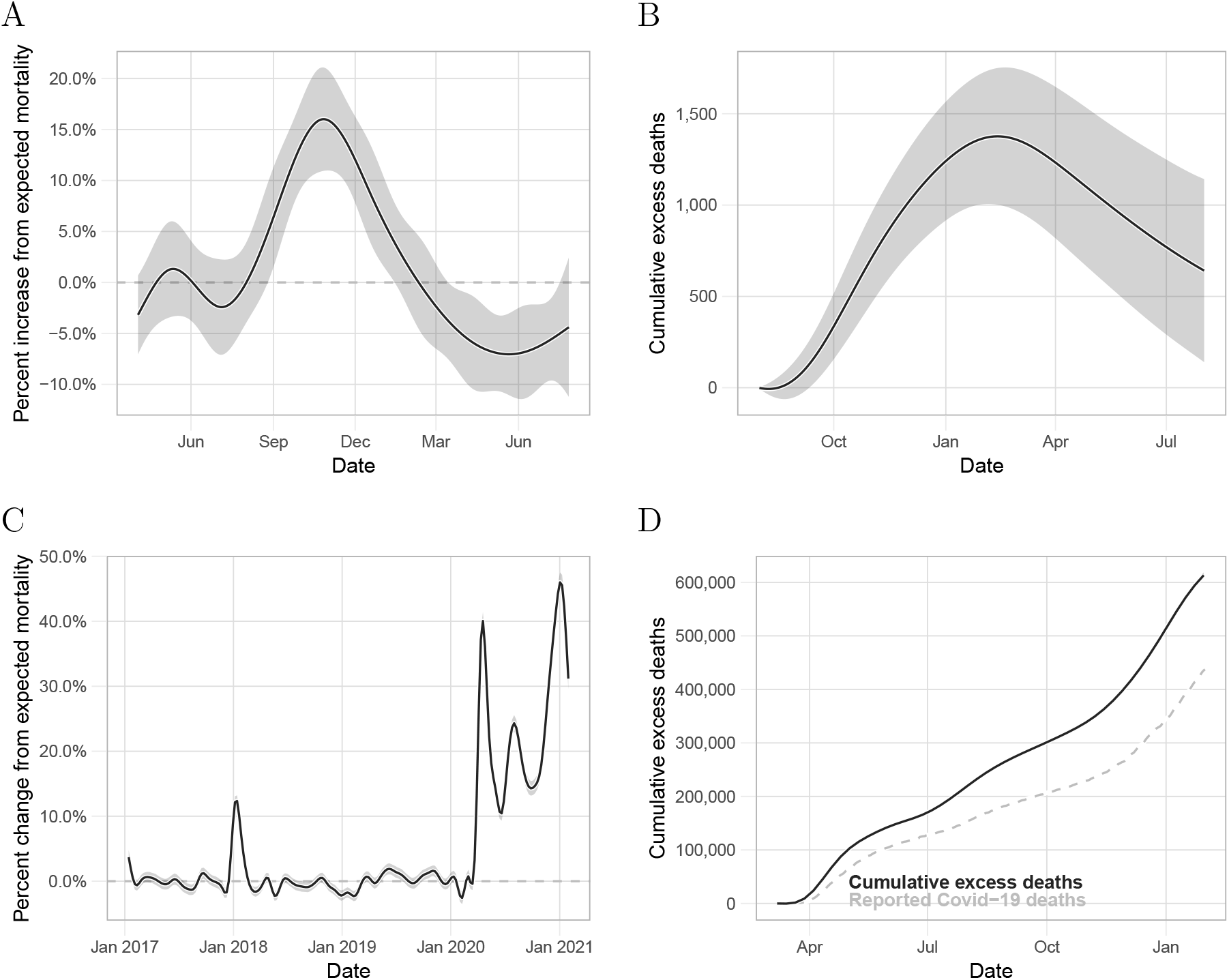
Detecting epidemics and outbreaks. A) Estimated percent change from expected mortality and point-wise 95% confidence interval for the period associated with the Chikungunya outbreak in Puerto Rico. B) Estimated cumulative excess deaths and point-wise confidence intervals for the period associated with the Chikungunya outbreak. C) Estimated percent change from expected mortality and point-wise 95% confidence interval for the United States from January 2017 to January 2021. D) Estimated cumulative excess deaths and point-wise confidence intervals for the United States during the period associated with the COVID-19 pandemic. The solid black curve is the cumulative COVID-19 related deaths reported by The New York Times.

As one further example, we implemented our approach to the US mortality data provided by the CDC to assess the effect of the COVID-19 pandemic. We aggregated the data from all states and then fit model (1) to obtain percent changes from expected mortality in the US (Figure 3C). To capture the rapid increase in mortality associated with COVID-19, we used 16 knots per year to estimate *f* (*t*). Because these are weekly counts, we fit the model assuming independent errors.

First, note that we capture the particularly bad 2017-18 flu. In 2020, we found an increase from average mortality associated with the COVID-19 pandemic that started in mid-March with a peak on the week ending on April 18, of 40% (95% CI: 38% to 42%). This was followed by a decrease lasting until mid-June. We then detected a second wave that peaked at the beginning of August with an increase from expected mortality of 24.3% (95% CI: 23.0% to 25.5%). In the subsequent weeks, excess mortality decreased until the end of Septembe, where then it increased until reaching its highest point since the beginning of the pandemic on the first week of 2021 with an increase from average mortality of 44.3% (95% CI: 42.8 to 45.8%). We found our cumulative excess deaths estimate to be larger than the cumulative COVID-19 deaths reported by the New York Times[16] (Figure 3D). Specifically, on 30 January 2021, the New York Times reported 435,441 COVID-19 deaths while our estimate was 604,400 (95% CI: 599,000 to 609,700) excess deaths in the United States, a difference of 168,959 excess deaths not directly accounted for by the reported COVID-19 deaths, indicated that not all deaths related to COVID-19 have been reported.

### Natural variability and correlated counts

Current approaches to excess mortality assume independent observations. Residual analysis of daily data, assuming independent errors demonstrates, the limitations of this assumption (eFigure 11). Assuming pairwise independence between a sequence of random variables, when in fact they are correlated, leads to downward bias of the standard error of the sum of the random variables. Therefore, if we incorrectly assume independence, the resulting standard errors will be an underestimate. This is particularly pernicious when computing standard errors for excess mortality estimates during long periods. To demonstrate this, we compared our method to a Poisson and over-dispersed Poisson model that assumes independent observations[7, 9, 11, 12, 13]. In the eAppendix we describe the details of our analysis. We found that the Poisson and over-dispersed Poisson model underestimates the underestimated standard errors. Conversely, modeling the correlation in the data, as done by our model, improved the standard error estimates (eFigure 12).

## Discussion

In this article we introduced a method and accompanying software that are useful for estimating excess mortality from daily counts. The engine of our approach is a statistical model that accounts for seasonality, secular trends, demographic changes, weekday effects, and natural variation in a unifying and parsimonious way. The biggest advantage of our method over previous ones is the characterization of indirect effects with smooth functions, which provides enough power to detect small effects that are long-lasting. Another advantage is that our model can be applied to daily data, which provides better resolution in detecting periods of concern. We demonstrated that current methods are not appropriate for daily counts because they do not model the correlation structure clearly observed in these data and thus yield underestimates of the standard errors. We were able to account for this by directly modeling the correlation using a mixed-effects model. This approach had the added advantage that we could incorporate the uncertainty introduced by the expected mortality estimate into our standard errors.

We note that using daily data often results in small counts and many zeros, in particular when data are stratified into specific demographic groups. We showed that by using smoothing our procedure produces accurate and unbiased estimates of the event effect even when the average number of deaths per day is as small as 0.10 per day. However, we advise users to perform exploratory analysis to assess model fit and provide tools to do this in our R package. Also, note that daily counts are not available in all jurisdictions. However, our approach is also applicable to lower resolution data such as weekly counts[25].

We demonstrated the utility of our approach by applying it to mortality data related to six hurricanes: Hugo, Georges, and Maria in Puerto Rico, Katrina in Louisiana, Sandy in New Jersey, and Irma in Florida. We found that the indirect effects of Hurricane Maria on Puerto Rico lasted several months after the storm, which is in accordance with previous findings[3, 4, 6, 5]. We also found a similar yet less severe pattern of indirect effects of Hurricane Georges on Puerto Rico. These event effects were much greater than those of Hurricanes Sandy and Irma on New Jersey and Florida, respectively. In contrast, the effects of Hurricane Katrina on Louisiana were much more direct, very possible due to the failure of the levees[26]. In agreement with previous findings, we further found that the 2014 Chikungunya epidemic resulted in substantial excess mortality in Puerto Rico[22]. These results suggest a lack of robustness in Puerto Rico’s health system. Finally, using our method we found stark discrepancies between our excess death estimates in the USA from March 2020 to January 2021, and the cumulative COVID-19 death toll reported by the New York Times. Specifically, we found three mortality waves coinciding with observed increases in cases[16].

When applying our methodology, users should be aware that the number of knots defining the splines changes the results. In general, we found that using 12 knots per year was appropriate for exploring the data and detecting unknown periods of excess mortality, while six knots per year was appropriate to characterize indirect effects for hurricanes and outbreaks. However, we highly recommend viewing diagnostic plots that help evaluate the model fit and sensitivity analysis to determine how these choices affect final summaries. Our software provides tools that facilitate this type of exploration. Users should also be aware that results depend on the population sizes *N*_*t*_ and that these are themselves estimates produced by government agencies. Finally, we note that it is not always apparent if a period showing increased mortality should be classified as natural variability (several consecutive large *ε*_*t*_s) or an event for which *f* (*t*) *>* 0. This choice will often have to be guided by context and expertise rather than data. Furthermore, it is important to consider that our method does not permit the decoupling of estimated effects from different events during the same period. For example, the estimated effect for Hurricane Maria in Puerto Rico ran into the winter of 2017-2018, during which time some US states were affected by an unusually severe flu season.

## Data Availability

We make our tools available through free and open source excessmort R package. All details of our analysis are available by examining the code which is made available on GitHub: https://github.com/RJNunez/excess-mortality-paper
The code for the R package implementing the methods is available on CRAN and on GitHub: https://github.com/rafalab/excessmort.

## Supplementary Material

### Statistical model

Here we provide more details on the model described by (1) and (2).

We assume *α*(*t*) is smooth enough that it can be represented by a smoothing cubic spline with one knot every seven years. If we have less than seven years of data, then *α*(*t*) is simply a linear function of time. To represent the seasonal trend we assumed a *harmonic model* :

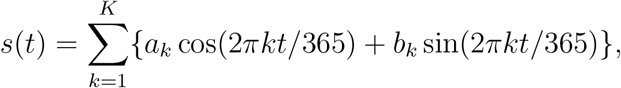

where the parameters ***a*** = [*a*_1_, …, *a*_*K*_] and ***b*** = [*b*_1_, …, *b*_*K*_] are estimated from data. Moreover, we model the weekday-specific effects using seven indicator variables and seven con-strained parameters:

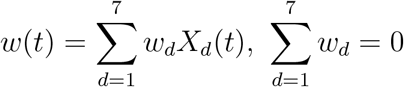

with *X*_*d*_(*t*) = 1 if day *t* is day of the week *d* and 0 otherwise. Exploratory data analysis demonstrated that this source of variability is pronounced in younger age groups that have higher death rates during the weekends. We note that if this source of variability is unaccounted for, the estimation of the parameters defining the error structure are affected. We further assumed the vector ***ε*** = [*ε*_1_, …, *ε*_*T*_]^*T*^ follows a truncated multivariate distribution with mean 1 (no change). To account for natural correlated variability we assumed ***ε*** had variance-covariance matrix, denoted with **Σ**, determined by an auto-regressive (AR) process of order *p*:

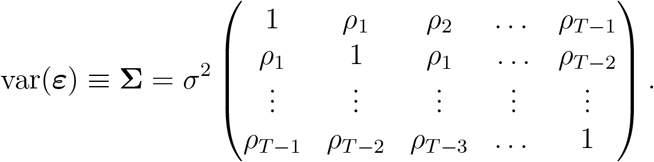

with *ρ*_1_, *… ρ*_*T −*1_ determined by the parameters of the AR(p) process. To assure 𝔼 (*Y*_*t*_ | *ε*_*t*_) *>* 0 we assumed that Pr(*ε*_*t*_ *>* 0) = 1. Note that the percent change in death rate due to natural variation, not accounted for seasonality and secular trends, observed in practice is exclusively between 20% smaller and 20% larger than mortality (0.8 *< ε*_*t*_ *<* 1.2), which consistent with this assumption.

### Estimating event effects

Here we describe the three-step approach that we use to estimate *f* (*t*) and standard error for these estimates. The general idea is to first estimate *μ*_*t*_ and **Σ** during periods with control periods, and then estimate the most interest component: *f* (*t*).

The first step is to estimate *α*(*t*), *s*(*t*), and *w*(*t*). To do this, we select a control period *I*control for which we know there were no natural disasters nor outbreaks and can assume *f* (*t*) = 0. Natural disasters and outbreaks are rare, hence, it should be possible to find such periods for most datasets. With this assumption in place model (1) reduces to:

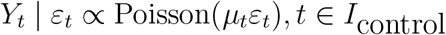

Note that if we have *N T/*365 years of daily data, given the choices described in the Tuning parameters section of the eAppendix, we are only fitting N/7 + 7 + 4 + 1 parameters to *T* data points. As an example, for seven years of data this translates into 2,556 data points and 13 parameters. As a result, we can obtain highly precise estimates 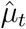 even in the presence of the extra dispersion introduced by *ε*_*t*_. We therefore fit a quasi-Poisson Generalized Linear Model (GLM) to regions in *I*control and estimate the expected value for *t ∉ I*control using the newly learned parameters. The quasi-Poisson assumption permits us to model the extra variability introduced by *ε*_*t*_.

In the second step we use the control region to estimate the variance-covariance matrix **Σ** as described in detail in the eAppendix. With this estimate in place we then use an iterative generalized least square procedure to estimate *f* (*t*) and its standard error. We use the Central Limit Theorem approximation to assume 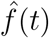 follows a normal distribution. As explained in the eAppendix, this standard error includes the variability introduced by the uncertainty in the estimate of the expected mortality rate 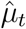.

### Estimating standard errors

The first step is to estimate the variance-covaiance matrix **Σ**. We use data in the control period to do this. First, let *r*_*t*_ be the observed percent change from expected mortality:

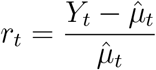

To propagate the uncertainty in the estimation process in the first step, we employ a first-order Taylor approximation of *r*_*t*_. Note that 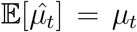 and 𝔼 [*Y*_*t*_] = *μ*_*t*_, where the latter equality holds by the law of total expectation. Then, we can approximate *r*_*t*_ around (*μ*_*t*_, *μ*_*t*_) and find that:

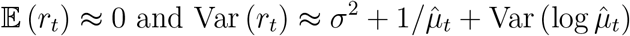

where we used the law of total variance to find Var(*Y*_*t*_). Intuitively, the terms in Var(*r*_*t*_) represent the variance added by *ε*, the Poisson variability, and the uncertainty from the first step, respectively. The above implies that the following random variable:

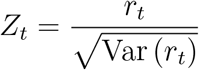

has the same correlation structure as *ε*_*t*_. We therefore use the Yule-Walker equations to estimate the AR process parameters from the observed *Z*_*t*_ within *I*control. To estimate *σ*^2^ we use:

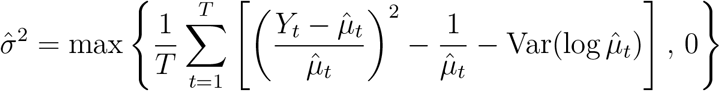

Using these we can form an estimate 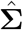.

In the third and final step we estimate *f* (*t*). Denote **r** = (*r*_1_ *…, r*_*T*_)^*T*^, **f** = (*f* (1), …, *f* (*T*))^*T*^ = **B*θ***, with **B** and ***θ*** the design matrix and parameters, respectively, that define the natural cubic spline and **D** a diagonal matrix with entries:

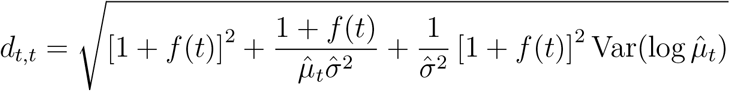

Then, we can use the fact that

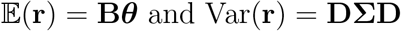

to obtain an unbiased estimate of **f** using generalized least squares:

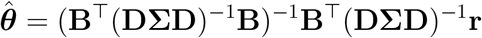

along with a variance estimate:

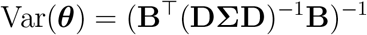

Since **DΣD** depends on the estimand **f**, we use an iterative procedure in which we plug-in the current estimate to compute the variance.

### Excess mortality estimate

To estiamte the standard error for 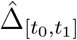, let ***B*** and ***θ*** be as they were defined above. Then, note that we can conveniently represent excess deaths at time *t* as *μ*_*t*_ *× f* (*t*) and define excess deaths for any time period [*t*_0_, *t*_1_] by just adding these up:

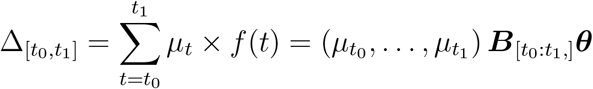

where 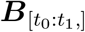 represents rows *t*_0_,…, *t*_1_ of matrix ***B***. We therefore estimate cumulative excess deaths for any interval *I* = [*t*_0_, *t*_1_] by adding the excess deaths for each day in *I*:

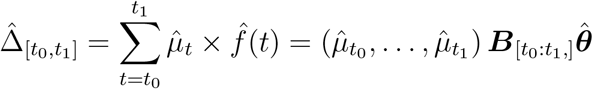

Where 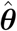 is the Maximum Likelihood estimate of ***θ***. We construct a 95% confidence interval using the CLT approximation to assume the sum is approximately normally distributed with variance given by:

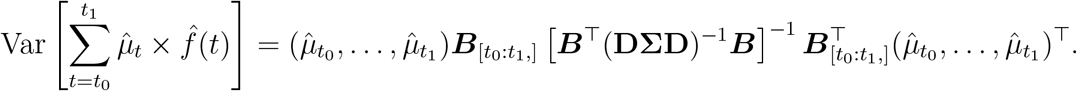

Note that our excess deaths estimate 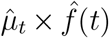 for time *t* is a smooth version of the estimate based on single time point: 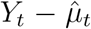. In fact, our estimate converges to 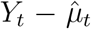 when the number of knots that define *f* is equal to the number of observations. The corresponding argument in our software package is the `knots_per_year` argument in the `excess_model` function.

## Mortality data stratified by demographic indicators

When mortality data is stratified by demographic indicators, such as age and sex, we can estimate event effect for each group. Note that we can then use this to estimate adjusted marginal effect. Specifically, if 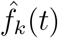 is the estimated effect at time *t* in group *k* we can define the adjusted overall-effect as event effect as a weighted sum of the group-specific effects:

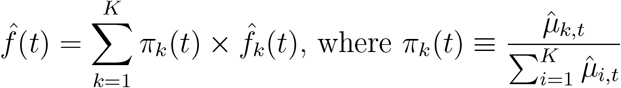

and 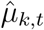 is the expected value of group *κ* at time *t*. As before, we can approximate Var 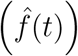 with a first-order Taylor approximation around (*f*_1_(*t*), *…, f*_*K*_(*t*), *μ*_1,*t*_, …, *μ*_*K,t*_) and find that:

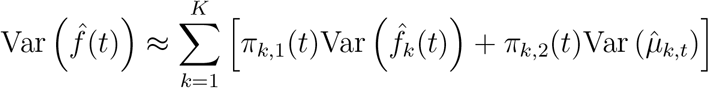

where

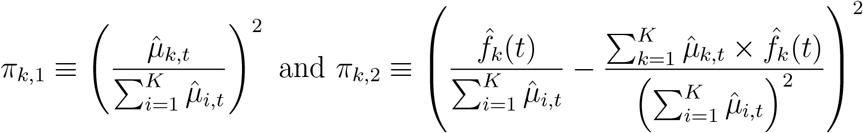

### Tuning parameters

Our approach included four fixed parameters that need to be defined before fitting the model to data. Here we motivate the default values we have set in the software implementation.

- We represent *α*(*t*) as a smoothing cubic spline with one knot every seven years. If we have less than seven years of data, then *α*(*t*) is simply a linear function of time. This choice is motivated by the observation that in many jurisdictions death rates have been slowly declining for the past decades.
- The seasonal trend is estimated by *s*(*t*), a periodic function with *K* harmonics. By computing and plotting the daily average count for each day of the year, for the 35 years of the Puerto Rico dataset, we noted that the pattern was not exactly sinusoidal (K=1) but that adding one harmonic (K=2) captured the extra complexity for most jurisdictions. We recommend generating this plot to guide the choice of *K* and we provide software in our package to do so.
- We can use our approach in an exploratory mode to search for periods in which *f* (*t*) ≠ 0. To do this, we use a flexible spline with 12 knots per year. This allows sharp increases to be captured. However, once we discover a period of increased death rates, we generate exploratory plots to determine if a smoother estimate is more appropriate. We used a spline with six knots per year for all hurricane effects and a discontinuity on landfall day.
- We set the degree of the AR process at seven. This choice was made by examining residual auto-correlation plots and assessments based on comparing observed and predicted standard errors for excess mortality estimates generated from periods for which we expected *f* (*t*) = 0. The software implementation permits the user to change this parameter or use AIC to determine the degree, as implemented in the R function ar.

### Population size estimates

We used yearly population estimates from the US Census, which correspond to the population size on July 1, to estimate daily population counts, *N*_*t*_, via linear interpolation. For days past the last value from the US Census, we assumed the population was constant and equal to the last day for which we had data. Yearly population estimates for Puerto Rico were obtained from the Puerto Rico Institute of Statistics (PRIS). Similarly, we estimated *N*_*t*_ via linear interpolation of the observed values. However, to account for population displacement after hurricane María [27, 28], we used estimates based on mobile phone records provided by Teralytics —a technology company that partners with telecommunication operators worldwide to assess human mobility[29]. Specifically, Teralytics provided daily population proportion estimates relative to a confidential baseline from May 2017, to April 2018. We multiplied these proportions by the 2017 mid-year population value from PRIS and generated smooth estimates with local regression. Finally, from the United Nations, we obtained age-specific population proportions in five year intervals for Puerto Rico from 1950 to 2020. Since we only have mortality data for Puerto Rico dating back to 1985, we generated daily demographic proportion estimates via linear interpolation of the five year interval values starting in 1985. Then, we computed age-specific population estimates by multiplying the demographic proportions times the aforementioned smooth population estimates.

### Simulation studies

To assess our procedure we conducted a Monte Carlo simulations. We designed simulation studies to mimic three scenarios 1) a natural disaster, 2) an infectious disease epidemic, and 3) a typical period with no events. The event effect, *f* (*t*), for the natural disaster scenario was defined by a strong direct effect followed by a slow decaying indirect effect, as seen after natural disasters. To achieve this, we fit model (2) to mortality data from Puerto Rico for all deceased 75 and older to periods with no known events. This provides estimates for 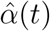, ŝ(*t*), and 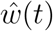 (eFigure 13) which we use to define *μ*_*t*_ for our simulations. We then fit model (1) to the period of July 20, 2017, to April 20, 2018, which includes Hurricane María, and used the resulting estimate 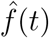 as the true *f* (*t*) for the first simulation study. To generate realistic effects *f* (*t*) for the infectious disease epidemic scenario, we fit model (1) from May 14, 2014, to February 14, 2015, which includes the Chikungunya outbreak in Puerto Rico, and use the resulting 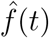 as the true *f* (*t*). Finally, we let *f* (*t*) = 0 for the entire period for the typical scenario simulation.

For natural variability we simulated serially correlated random variables {*ε*_1_, …, *ε*_*T*_} centered at one with an AR process of degree two and coefficients obtained using the Yule-Walker equations (see eAppendix for details). We set the standard deviation of *ε*_*t*_ to 0.05 and used data from January 1, 2002 to December 31, 2013, a contiguous time period in which no known events occurred, to estimate the coefficients. With these parameters in place we then generated *B* = 100, 000 simulated datasets following model (1):

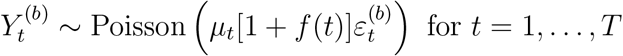

for *b* = 1, …, *B*. Note that 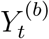 refers to the simulated number of deaths at time *t* for simulation *b*. In eFigure 1, the solid-red curves represent the true curve or true standard error, whereas the dashed-black curves correspond to our estimates. We find that our method consistently estimates the true curve precisely under all three scenarios (eFigures 1A-C). Our estimated standard error also estimates the true standard error precisely (eFigures 1D-F).

### Small death counts

The smoothing approach we employ improves power over methods based on measurements from a single day, particularly in situations where the signal to noise ratio is low. This implies that our method can be useful even when the death counts per time unit are small. We conducted a simulation study to determine the lower limit for deaths per day rates that our approach can handle. Specifically, we simulated data using the same approach we described above, but we normalized the counts such that the average deaths per day was *γ* = *{*0.05, 0.10, 0.50, 1.00*}* across the simulated data. Specifically, we use:

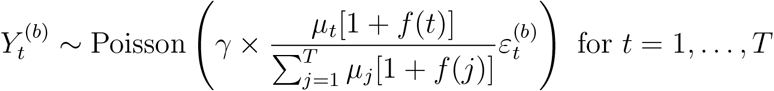

The simulation demonstrated that for daily count rates above 0.10 our model performs well (eTable 1, eFigures 2 & 3). Finally, we note that for the demographics groups described in Section and used in Sections and, the lowest counts per day rate was 0.30.

### Amount of training data

The first step of our approach is to estimate the expected counts *μ*_*t*_ for each time *t* using data in the control region. To assess the impact of the length of this control baseline period in our estimation procedure, we employed the same simulation scheme we described above, but we limited the length of the control region to 2, 4, 6, and 8 years. We found that results were practically equivalent for all training periods (eTable 4, eFigures 4 & 5)

### False Discovery Rates and Power Analysis

To study the false discovery rate of our procedure we again repeated the simulation used in Section using the null model *f* (*t*) = 0. For each simulated dataset, we fit our model with 6 and 12 knots per-year. We also considered the saturated model that results in the estimate 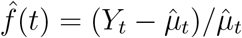 For each model fit, we then searched for regions and recorded recorded all intervals for which 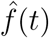 was statistically different from 0 and the length of this interval. We then considered several period length requirements and reported the number of detected false events per year.

We repeated the simulation above but this time for a case in which *f* (*t*) *>* 0. Specifically, we defined *f* using the Tukey tri-weight function:

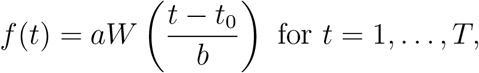

where *t*_0_ is the day where the effect is highest, *a* is the peak effect, 2*b* is the length of the effect in days, and *W* (*u*) = (1 − |*u*| ^3^)^3^, for |*u*|≤ 1. For this simulation we set *a* = 0.20 and *b* = 45, hence the period of indirect effect lasted 90 days. Similar to above, for each period length requirement, we recorded the proportion of years where one of our detected intervals had a non-null intersection with the interval for which *f* (*t*) *>* 0, specifically [*t*_0_ − *b, t*_0_ + *b*]. We reported the proportion of years for which the event of concern was detected.

We found that our method greatly improved sensitivity over the saturated model approach without much loss of specificity (eTables 2 & 3). If we require to see at least 5, 10, or 30 days above the threshold with a 12 knot smoother, then we see a false event every 2, 3, and 33 years, respectively, As described in the Methods section, we can also control specificity and sensitivity by increasing the confidence level.

### Natural variability and correlated counts

To demonstrate the perilous effects of incorrectly assuming independence between counts, when in fact they are correlated, we compared our method to a Poisson and over-dispersed Poisson model that assumes independent observations. We fit each of the three models to the Puerto Rico data for individuals 75 and over. Then, we randomly selected 100 intervals of sizes *L* = 10, 50, and 100 days from periods with no events, and computed the total number of deaths in each interval *S*_*l*_ = Σ_*t∈L*_ *Y*_*t*_. We then used the fitted models to estimate the expected value 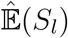 and standard error 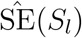. If the expected value and the variance are estimated correctly, the Central Limit Theorem predicts that the statistic 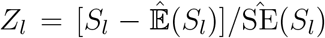 should have an approximate Gaussian distribution with mean zero and variance one. We found that the Poisson and over-dispersed Poisson model underestimates the SE(*S*_*l*_) resulting in larger than expected values of |*Z*_*l*_|.

## Supplementary Tables

**Supplementary Table 1:** Assessment of the performance of our model when the rate of deaths per day is small. The first column shows the rate of deaths per day in each simulation. The second column shows the median, over time, of the absolute value of the bias of our estimate of the *event effect*. The third column shows the median, over time, of the absolute value of the bias of our estimate of the standard error of 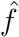. The fourth column shows the median, over time, root mean squared error of our estimate of the standard error of 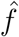.

| Counts<br>per day | Estimate<br>median bias | SE estimate<br>median bias | SE estimate<br>median RMSE |
| --- | --- | --- | --- |
| 1.00 | 0.0004 | 0.0003 | 0.0082 |
| 0.50 | 0.0005 | 0.0013 | 0.0156 |
| 0.10 | 0.0030 | 0.0582 | 0.1073 |
| 0.05 | 0.0153 | 0.1277 | 0.2324 |

**Supplementary Table 2:** False discovery rates based on a simulation study where *f* (*t*) = 0, for all *t*. The results are shown as the rate of false events detected per year, out of the 100,000 simulated years. The first column shows the amount of smoothness. The second column shows results for all detected regions. Column 3 through 7 shows results for regions of length 3, 5, 10, 30, 60 or larger, respectively.

| knots | $\geq 1$ day | $\geq 3$ days | $\geq 5$ days | $\geq 10$ days | $\geq 1$ month | $\geq 2$ months |
| --- | --- | --- | --- | --- | --- | --- |
| 6 | 0.327 | 0.324 | 0.318 | 0.295 | 0.140 | 0.011 |
| 12 | 0.561 | 0.549 | 0.525 | 0.424 | 0.038 | 0.000 |
| Saturated* | 4.659 | 0.001 | 0.000 | 0.000 | 0.000 | 0.000 |

**Supplementary Table 3:** Power analysis based on a simulation study where *f* (*t*) *>* 0 for an interval of 90 days. The results are shown as the rate of years in which we correctly detect the event of concern, out of the 100,000 simulated years. The first column shows the amount of smoothness. The second column shows results for all detected regions. Column 3 through 7 shows results for regions of length 3, 5, 10, 30, 60 or larger, respectively.

| knots | $\geq 1$ day | $\geq 3$ days | $\geq 5$ days | $\geq 10$ days | $\geq 1$ month | $\geq 2$ months |
| --- | --- | --- | --- | --- | --- | --- |
| 6 | 1.000 | 1.000 | 1.000 | 1.000 | 1.000 | 0.999 |
| 12 | 1.000 | 1.000 | 1.000 | 1.000 | 1.000 | 0.762 |
| Saturated* | 1.000 | 0.119 | 0.003 | 0.000 | 0.000 | 0.000 |

**Supplementary Table 4:** Assessment of the effect of the length of the control period used to estimate the expected counts. The first column shows the rate of deaths per day in each simulation. The second column shows the median, over time, of the absolute value of the bias of our estimate of the *event effect*. The third column shows the median, over time, of the absolute value of the bias of our estimate of the standard error of 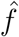. The fourth column shows the median, over time, root mean squared error of our estimate of the standard error of 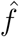.

| Length of the control region | Estimate median bias | SE estimate median bias | SE estimate median RMSE |
| --- | --- | --- | --- |
| 8 | 0.0001 | 0.0035 | 0.0041 |
| 6 | 0.0001 | 0.0023 | 0.0032 |
| 4 | 0.0001 | 0.0031 | 0.0043 |
| 2 | 0.0001 | 0.0020 | 0.0040 |

## Supplementary Figures

**Supplementary Figure 1:**
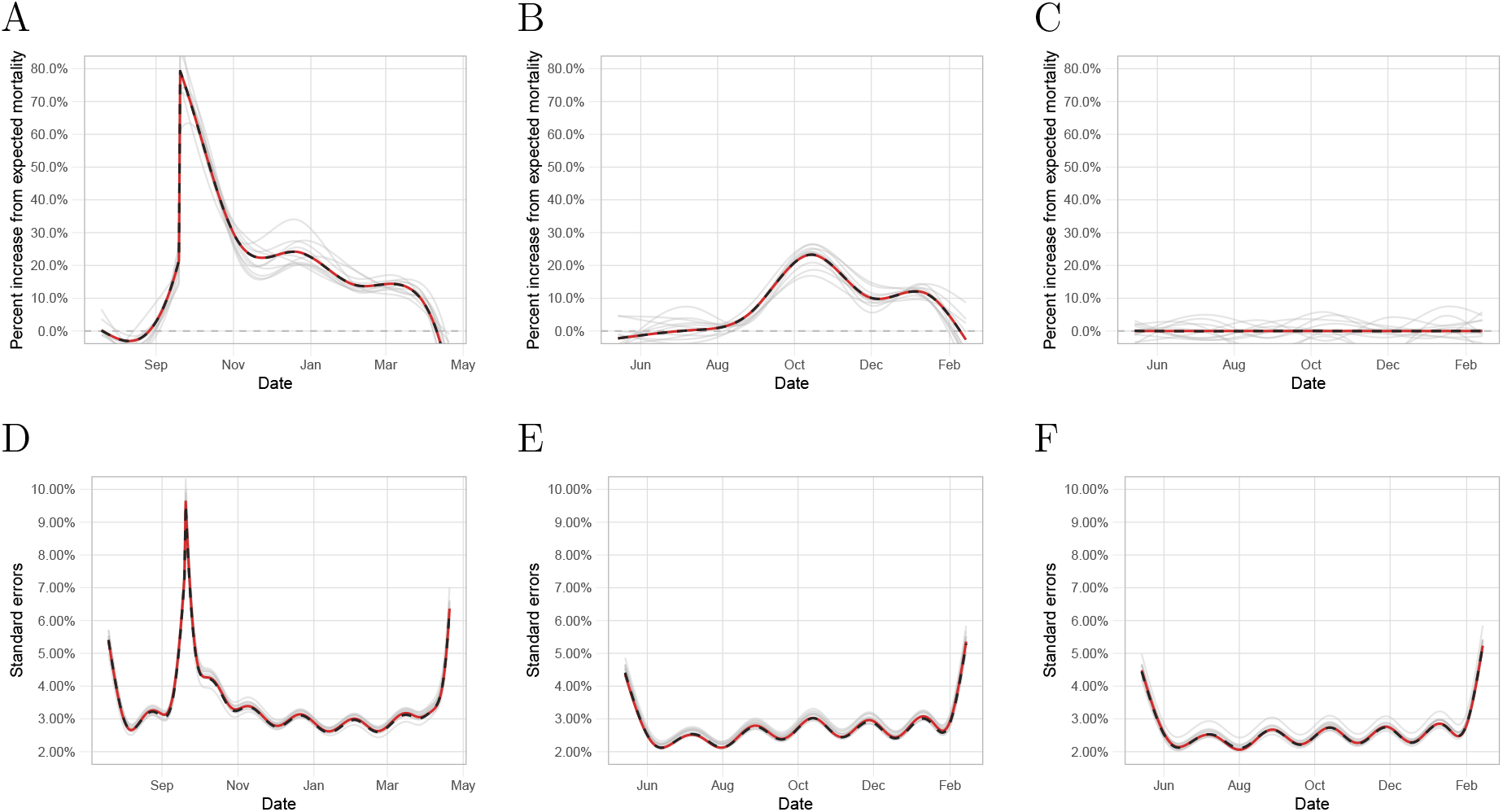
Assessment of our procedure based on a simulation study. A) The solid-red curve represents the true event effect, *f* (*t*), for the natural disaster scenario. The dashed-black curve is the Monte Carlo approximation of 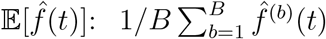. The grey curves are a random sample of ten event effects, 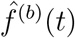. B) As A) but for the infectious disease epidemic scenario. C) As A) but for the typical scenario. D) The solid-red curve represents the Monte Carlo approximation of the standard error 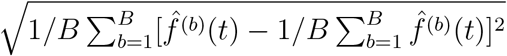. The dashed-black curve represents the average of the standard errors across all simulations: 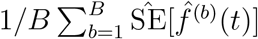. The grey curves represent a random sample of ten such standard errors: 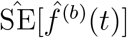. E) As D) but for the infectious disease epidemic scenario. F) as D) but for the typical period scenario.

**Supplementary Figure 2:**
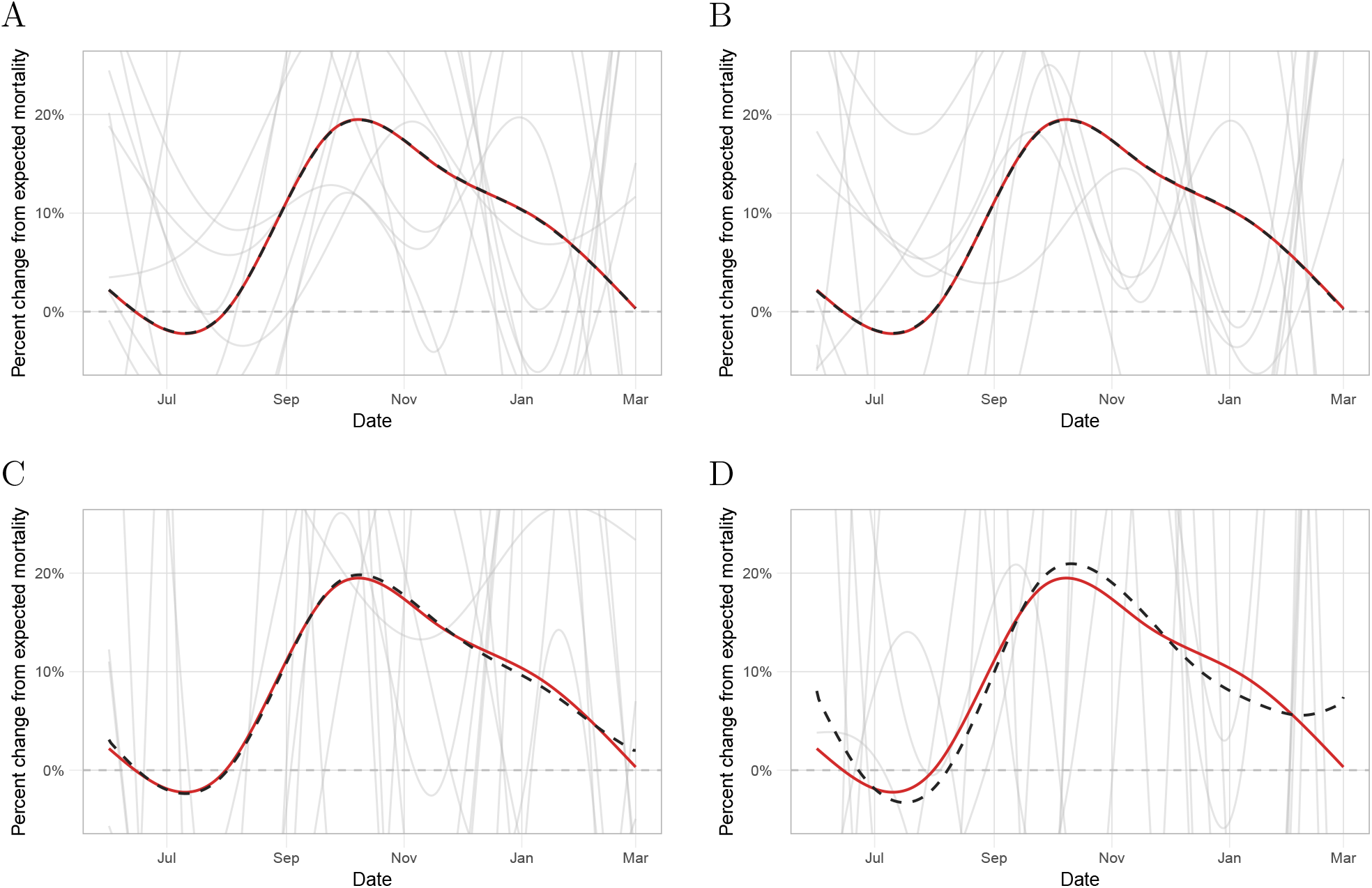
Assessment of our estimate of the *event effect* based on a simulation study where the number of deaths per day was small. A) The average number of deaths per day was set to 1. The solid-red curve represents the true *event effect, f* (*t*). The dashed-black curve is the Monte Carlo approximation of 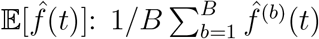. The grey curves are a random sample of ten *event effects*, 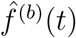. B) As A) but for a scenario where the average number of deaths per day was set to 0.50. C) As A) but for a scenario where the average number of deaths per day was set to 0.10. D) As A) but for a scenario where the average number of deaths per day was set to 0.05.

**Supplementary Figure 3:**
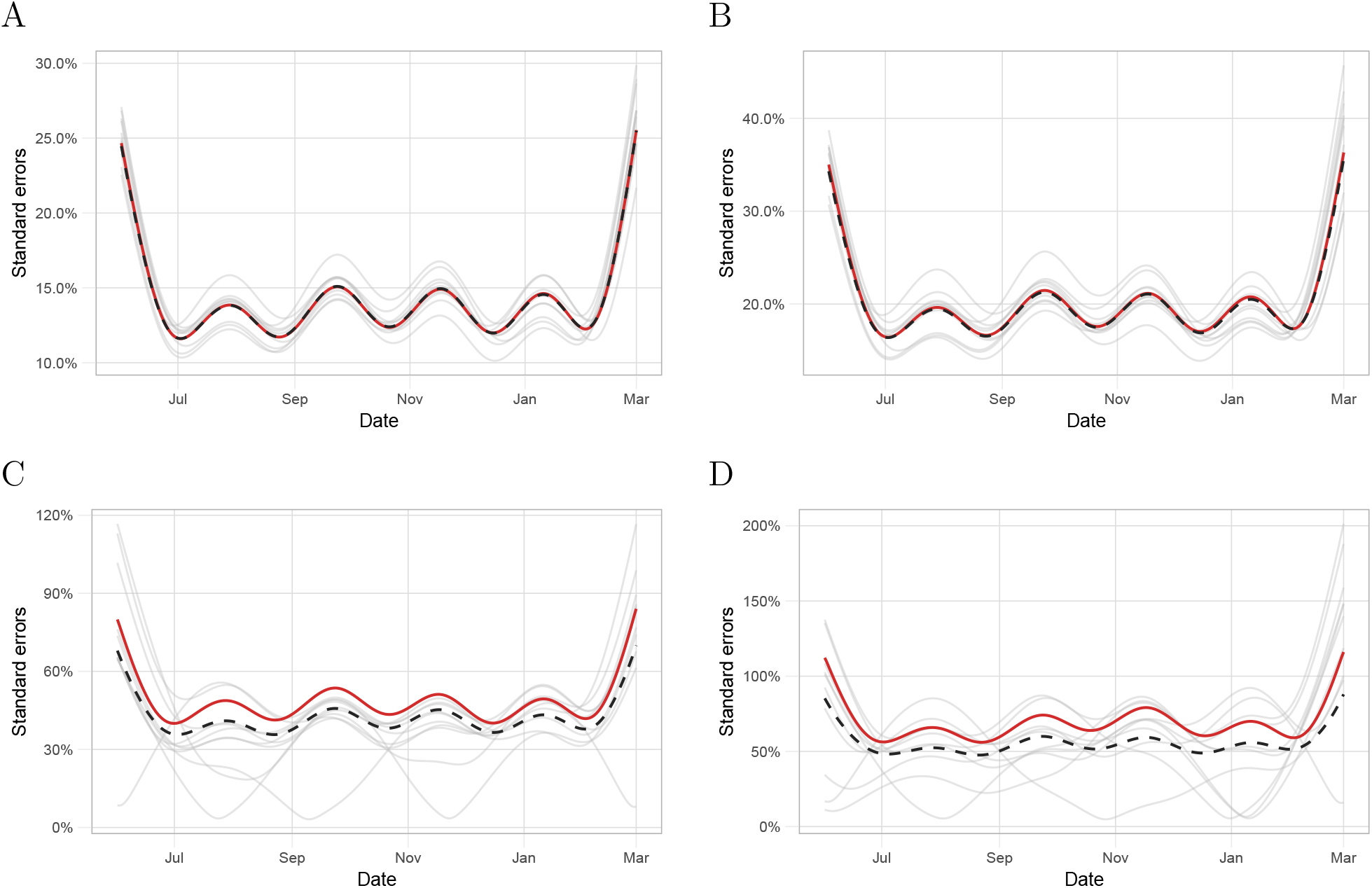
Assessment of our estimate of the standard error of the *event effect* based on a simulation study where the number of deaths per day was small. A) The average number of deaths per day was set to 1. The solid-red curve represents the Monte Carlo approximation of the standard error 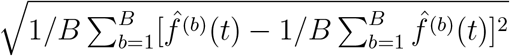. The dashed-black curve represents the average of the standard errors across all simulations: 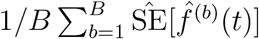. The grey curves represent a random sample of ten such standard errors: 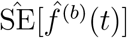. B) As A) but for a scenario where the average number of deaths per day was set to 0.50. C) As A) but for a scenario where the average number of deaths per day was set to 0.10. D) As A) but for a scenario where the average number of deaths per day was set to 0.05.

**Supplementary Figure 4:**
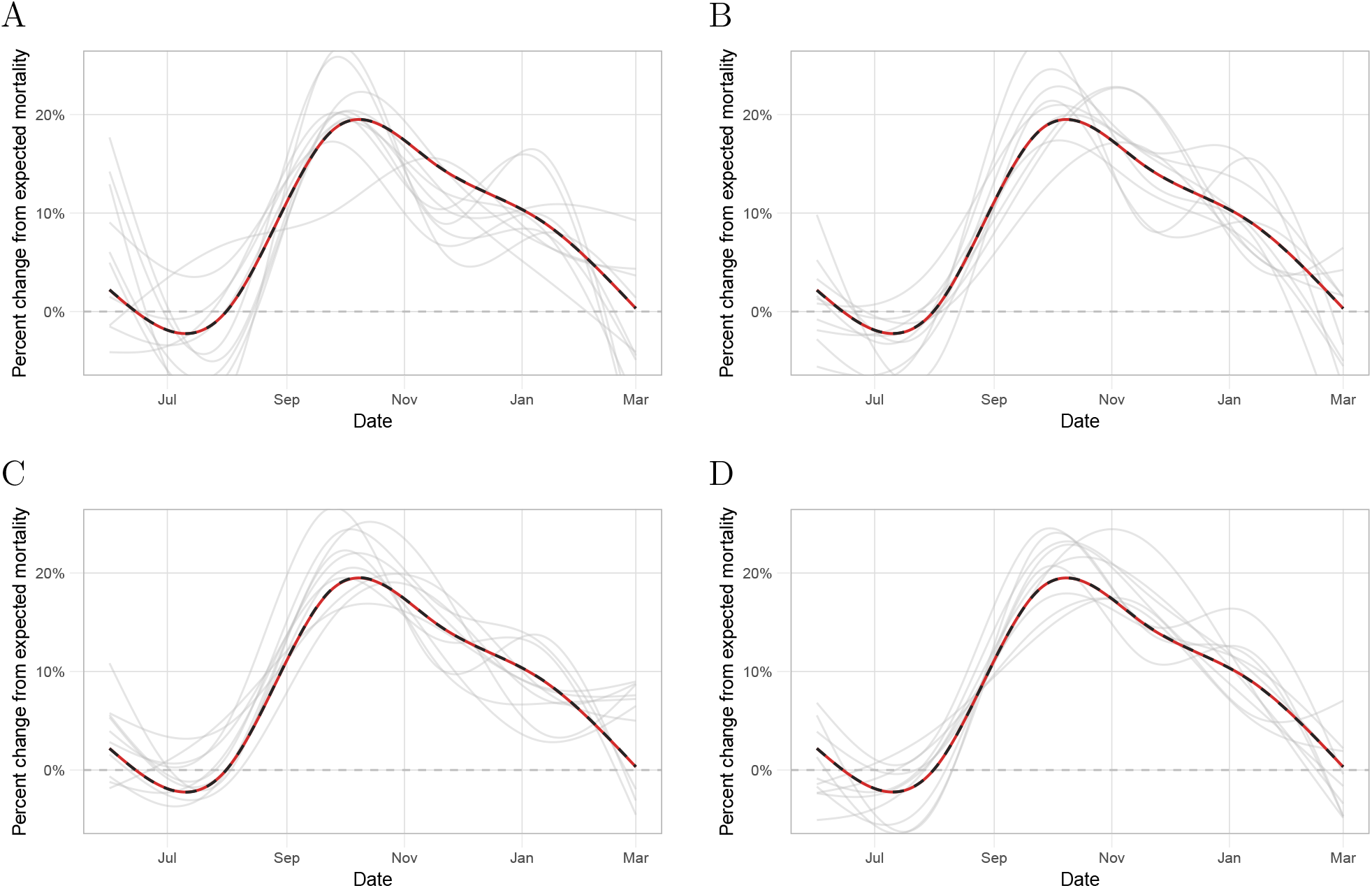
Assessment of our estimate of the *event effect* based on a simulation study where we varied the length of the control region. A) The length of the control region was 8 years. The solid-red curve represents the true *event effect, f* (*t*). The dashed-black curve is the Monte Carlo approximation of 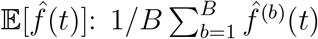. The grey curves are a random sample of ten *event effects*, 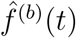. B) As A) but for a scenario where the length of the control region was 6 years. C) As A) but for a scenario where the length of the control region was 4 years. D) As A) but for a scenario where the length of the control region was 2 years.

**Supplementary Figure 5:**
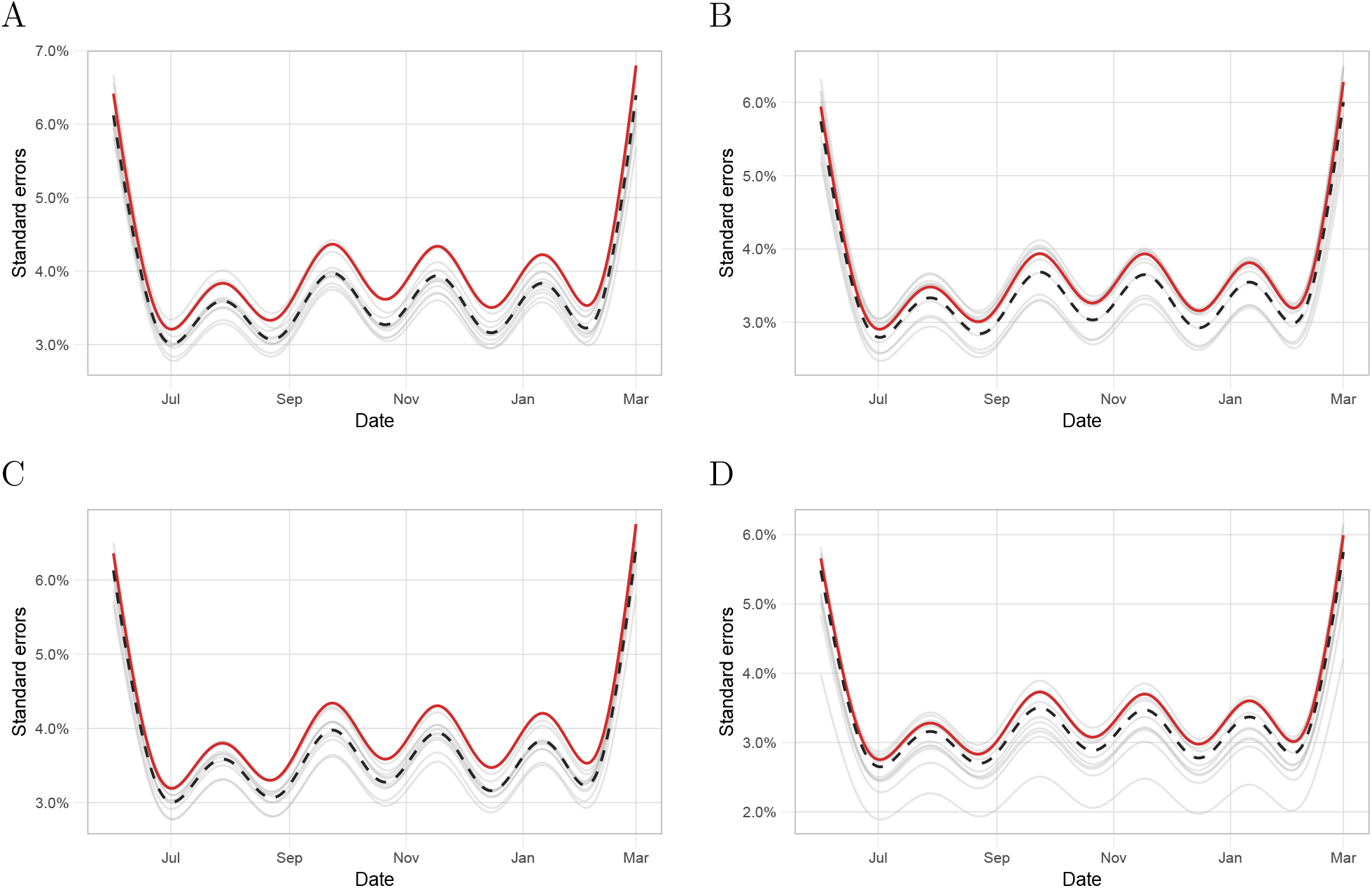
Assessment of our estimate of the standard error of the *event effect* based on a simulation study where we varied the length of the control region. A) The length of the control region was 8 years. The solid-red curve represents the Monte Carlo approximation of the standard error 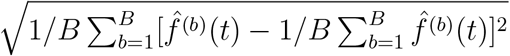. The dashed-black curve represents the average of the standard errors across all simulations: 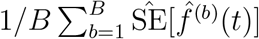. The grey curves represent a random sample of ten such standard errors: 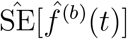. B) As A) but for a scenario where the length of the control region was 6 years. C) As A) but for a scenario where the length of the control region was 4 years. D) As A) but for a scenario where the length of the control region was 2 years.

**Supplementary Figure 6:**
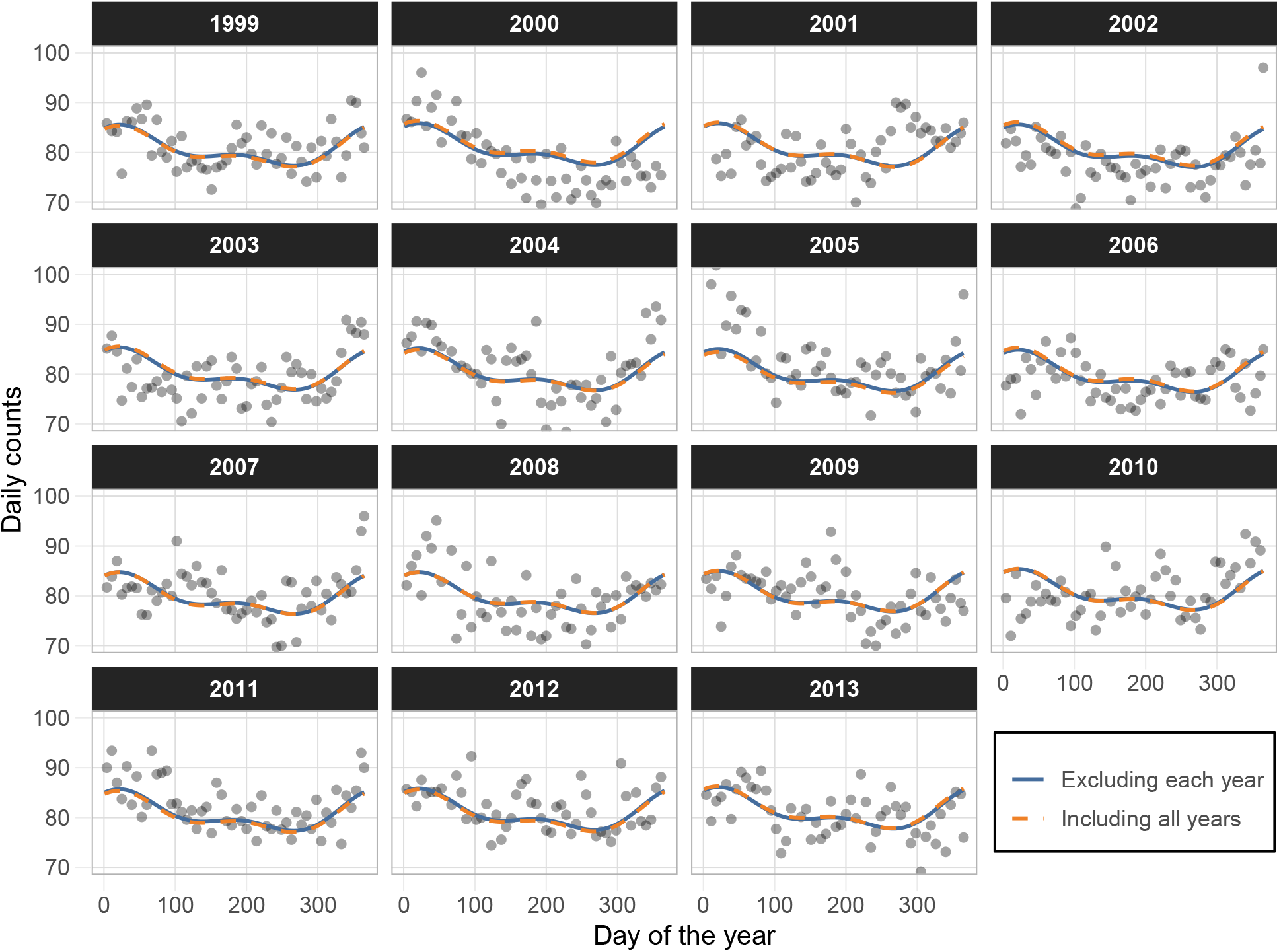
Cross-validation study to assess the performance of our mean model. For years 1999 to 2013 in Puerto Rico we removed each year, one by one, estimated *μ*_*t*_ without that year and compared it to the estimate obtained when including that year in the analysis. The title of each pane represents the year that was removed. The points are the average deaths for every week, the solid-blue curve is the expected value when excluding each year, and the dashed-orange curve is the expected value when we did not exclude each year.

**Supplementary Figure 7:**
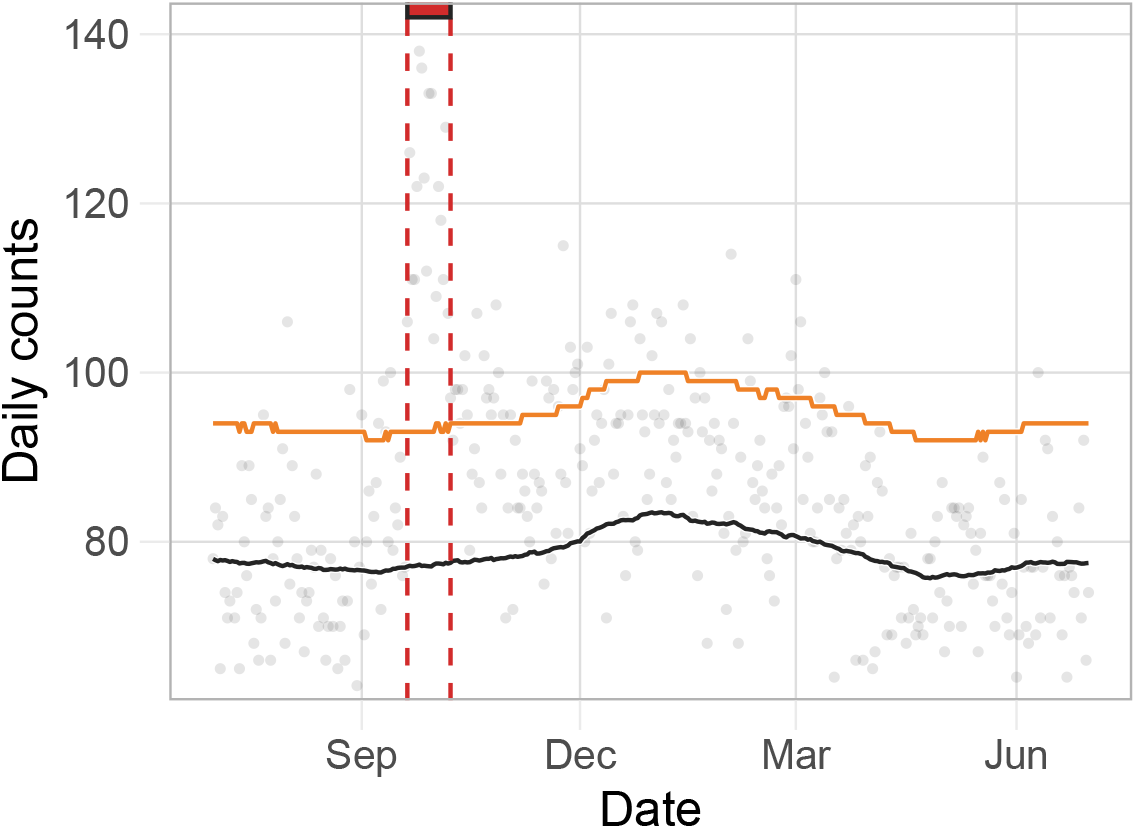
Farrington model fit to daily Puerto Rico data for a period that includes the landfall of Hurricane Maria. Gray points represent daily deaths counts. The black and the orange curves are the expected number of daily counts and the threshold for significant excess deaths, respectively, as defined by the Farrington algorithm. The red rectangle denotes the number of consecutive days with excess deaths since the landfall of Hurricane Maria as determined by the Farrington algorithm.

**Supplementary Figure 8:**
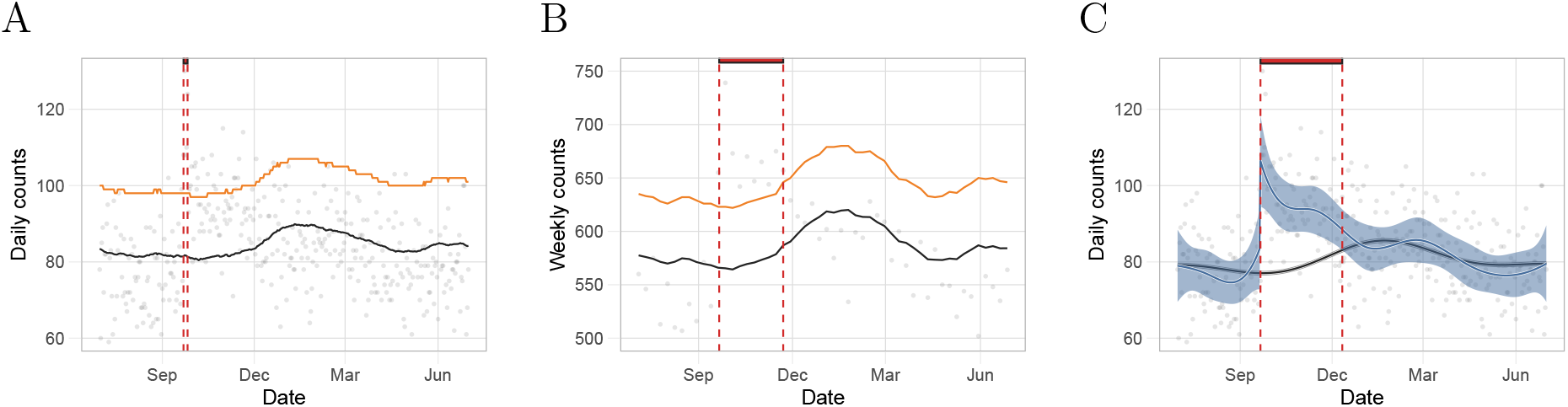
Comparison between the Farrington model and our method based on estimates for Puerto Rico from a period including Hurricane Georges. A) Gray points represent daily deaths counts. The black and the orange curves are the expected number of daily counts and the threshold for significant excess deaths, respectively, as defined by the Farrington algorithm. The red rectangle denotes the number of consecutive days with excess deaths since the landfall of Hurricane Maria as determined by the Farrington algorithm. B) Gray points represent weekly death counts. The black and the orange curves are the expected number of daily counts and the threshold for significant excess deaths, respectively, as defined by the Farrington algorithm. The red rectangle denotes the number of consecutive week with excess deaths since the landfall of Hurricane Maria as determined by the Farrington algorithm. C) Gray points represent daily death counts. The black curve is the estimated expected counts based on our method and the blue curve represents the *event effect* estimate, 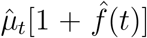. The black and blue ribbons are point-wise 95% confidence intervals for the expected counts and *event effect*, respectively. Finally, the red rectangle is as in B) but for our method.

**Supplementary Figure 9:**
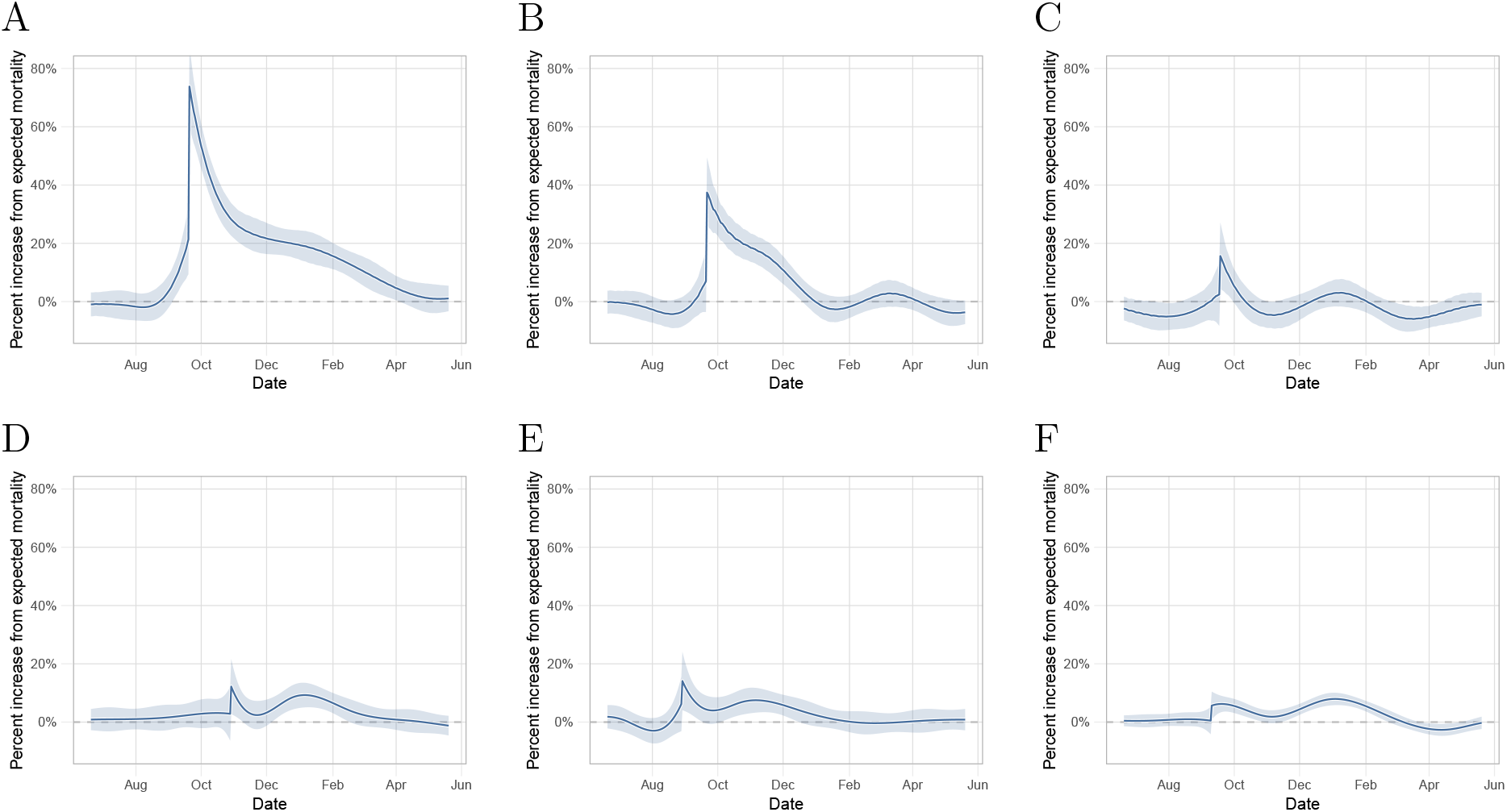
Estimated hurricane effects as percent increase over expected mortality for the six hurricanes. A) *Event effect* of Hurricane Maria in Puerto Rico. The blue curve and ribbon represent the *event effect*, 100 *× f* (*t*), and corresponding point-wise 95% confidence intervals. B) As A) but for Hurricane Georges in Puerto Rico. C) As A) but for Hurricane Hugo in Puerto Rico. D) As A) but for Hurricane Sandy in New Jersey. E) As A) but for Hurricane Katrina in Louisiana. F) As A) but for Hurricane Irma in Florida.

**Supplementary Figure 10:**
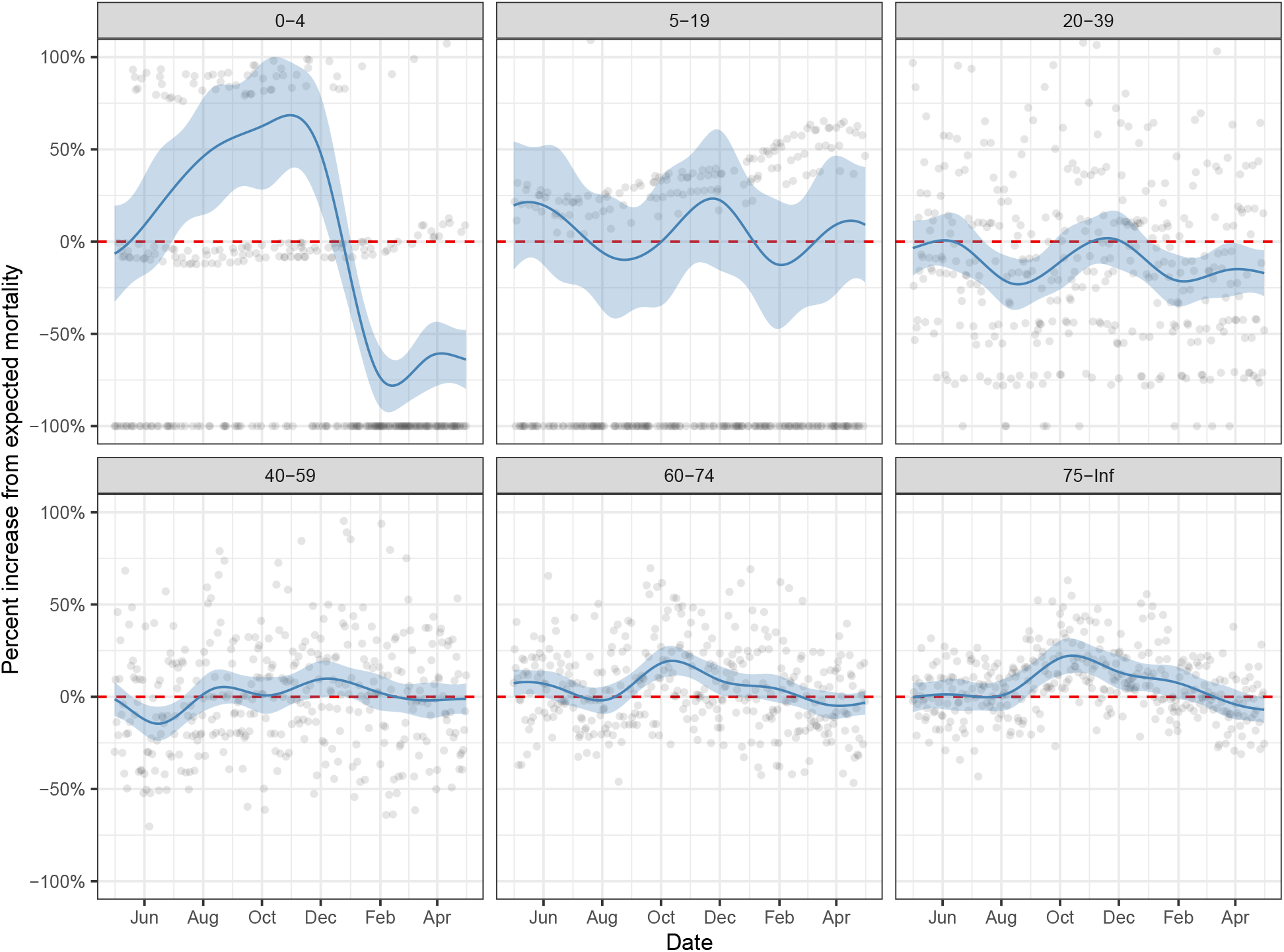
Estimated effects as percent increase over expected mortality during the Chikungunya epidemic for different age groups. The grey data points correspond to observed daily percent changes from expected mortality. The blue curve and ribbon represent the event effect and its point-wise 95% confidence interval.

**Supplementary Figure 11:**
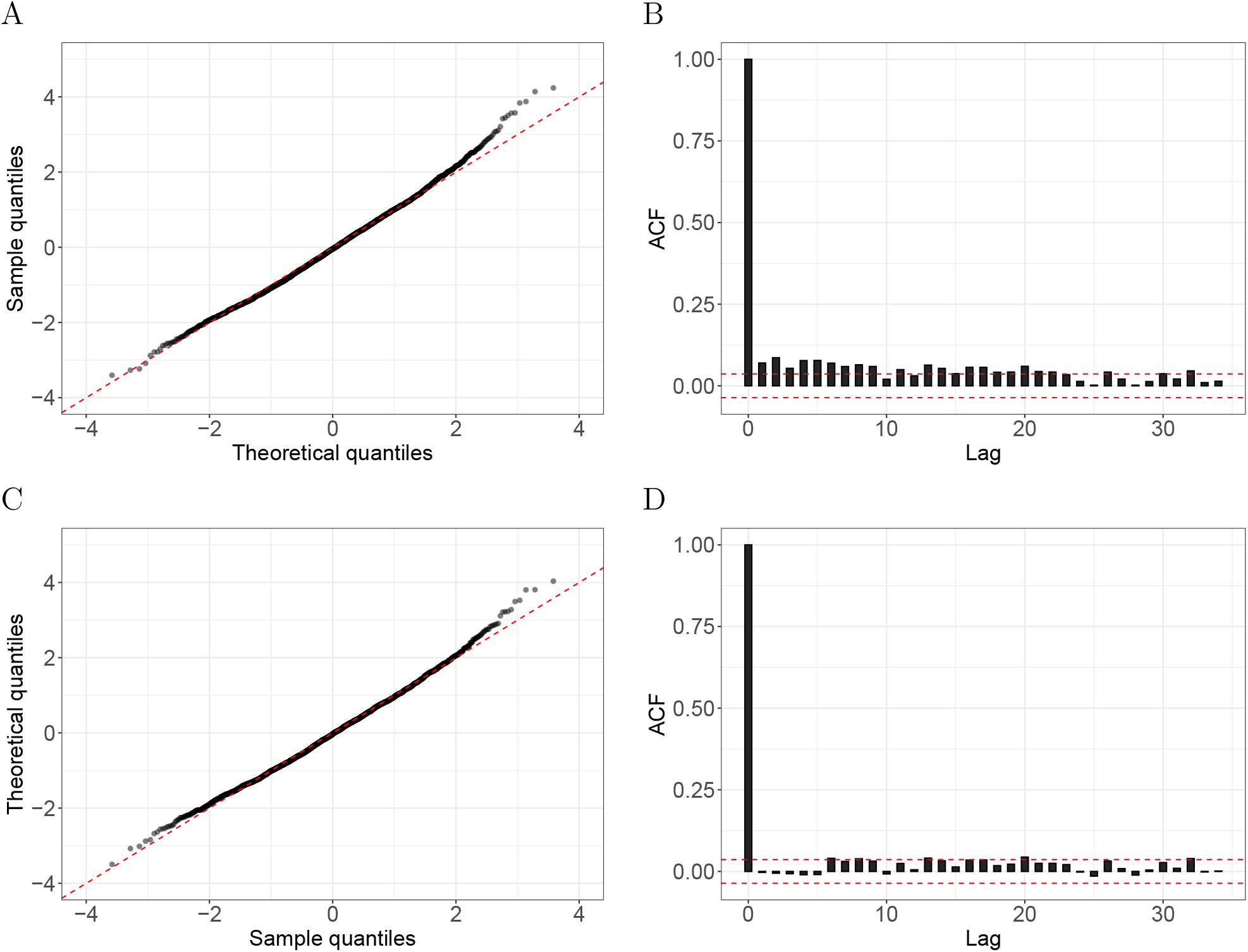
Evidence of correlated errors. A) A Poisson GLM was fitted to Puerto Rico daily death counts of individuals 75 years and older from an interval with no known natural disasters or outbreaks (Jan 1, 2006 to Dec 31, 2013). The plot shows the Pearson residual quantiles versus theoretical quantiles from the normal distribution. One can see that the tail of the empirical data are larger than the theoretical values. B) The sample autocorrelation function for these Pearson residuals with the red-dash lines represent a 95% confidence interval centered at zero. C) As A) for residuals that were adjusted for the correlation in the data based on an estimate of the covariance matrix. D) As B) for residuals that were adjusted for the correlation in the data based on an estimate of the covariance matrix.

**Supplementary Figure 12:**
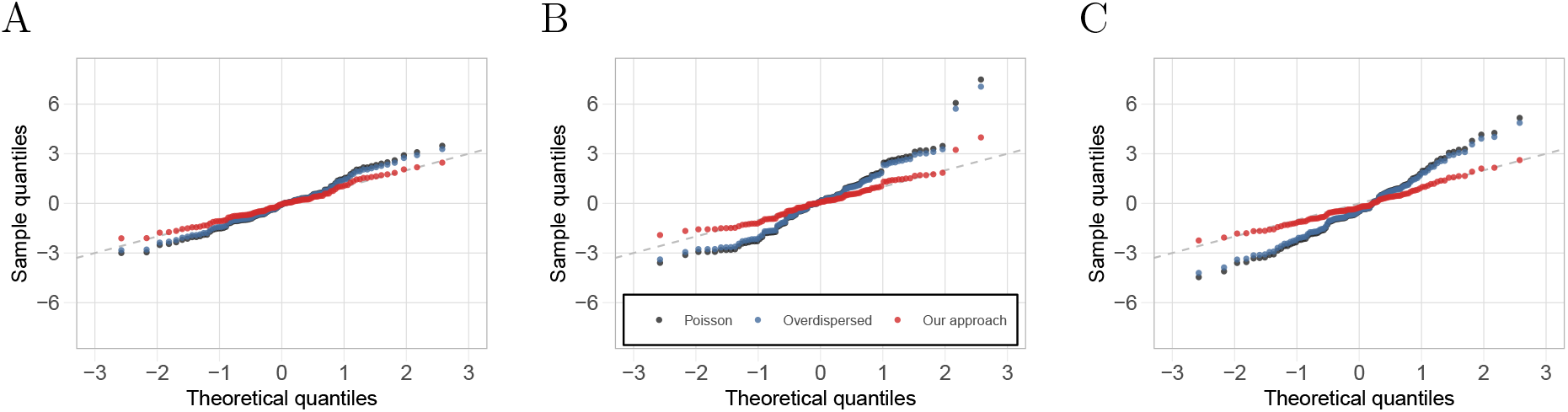
Accounting for correlation in the error structure improves uncertainty estimates. A) Wald-statistics versus theoretical quantiles for the standard normal distribution for excess deaths estimated based on 10 days in the control interval. B) As A) but for 50 day intervals. C) As A) but for 100 day intervals.

**Supplementary Figure 13:**
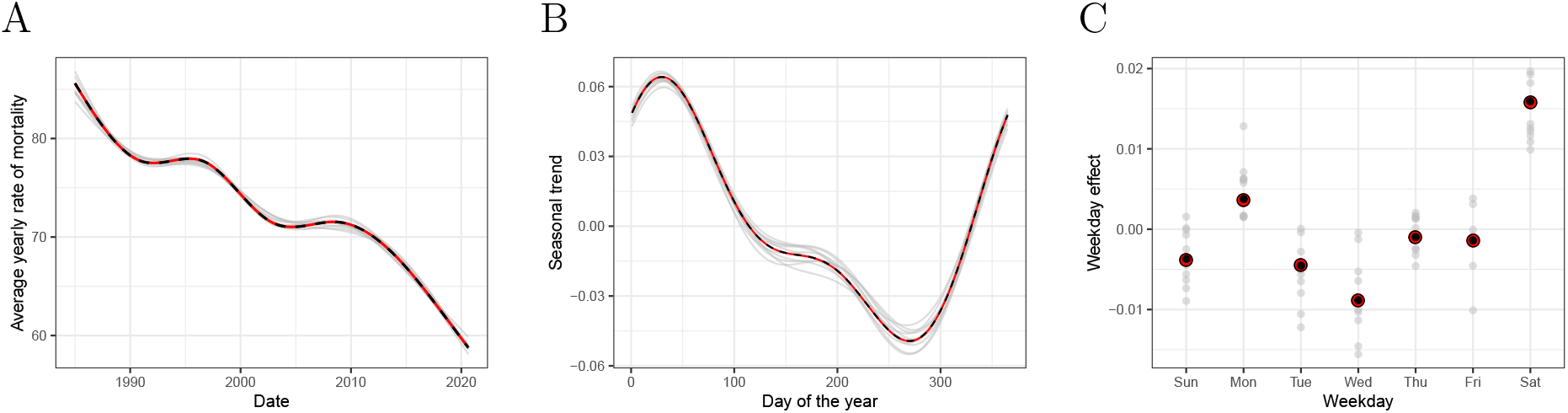
Assessment of the mean model based on a simulation study. A) The solid-red curve represents the true *α*(*t*), the dashed-black curve is the Monte Carlo approximation of the expected value of this estimate: 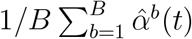, and the grey curves are a random sample of 10 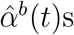. B) As A) but for *s*(*t*). C) The red points represent the true *w*(*t*), the black points are the Monte Carlo approximation of the expected value of these estimates: 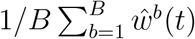, and the grey points are a random sample of 10 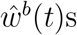.

## References

[1] US Department of Health, Human Services, National Center for Health Statistics, et al. A reference guide for certification of deaths in the event of a natural, human-induced, or chemical/radiological disaster. Hyattsville, Maryland USA: CDC, 2017.

[2] Esteban Ortiz-Ospina Max Roser, Hannah Ritchie and Joe Hasell. Coronavirus pan-demic (covid-19). Our World in Data, 2020. https://ourworldindata.org/coronavirus.

[3] Carlos Santos-Burgoa, John Sandberg, Erick Suárez, Ann Goldman-Hawes, Scott Zeger, Alejandra Garcia-Meza, Cynthia M Pérez, Noel Estrada-Merly, Uriyoan Colón-Ramos, Cruz María Nazario, et al. Differential and persistent risk of excess mortality from hurricane maria in puerto rico: a time-series analysis. The Lancet Planetary Health, 2 (11):e478–e488, 2018.

[4] Roberto Rivera and Wolfgang Rolke. Modeling excess deaths after a natural disaster with application to hurricane maria. Statistics in medicine, 38(23):4545–4554, 2019.

[5] Raul Cruz-Cano and Erin L Mead. Causes of excess deaths in puerto rico after hurricane maria: a time-series estimation. American journal of public health, 109(7):1050–1052, 2019.

[6] Alexis R Santos-Lozada and Jeffrey T Howard. Use of death counts from vital statistics to calculate excess deaths in puerto rico following hurricane maria. Jama, 320(14): 1491–1493, 2018.

[7] Daniel M Weinberger, Jenny Chen, Ted Cohen, Forrest W Crawford, Farzad Mostashari, Don Olson, Virginia E Pitzer, Nicholas G Reich, Marcus Russi, Lone Simonsen, et al. Estimation of excess deaths associated with the covid-19 pandemic in the united states, march to may 2020. JAMA Internal Medicine, 180(10):1336–1344, 2020.

[8] Gonzalo E. Mena, Pamela P. Martinez, Ayesha S. Mahmud, Pablo A. Marquet, Caroline O. Buckee, and Mauricio Santillana. Socioeconomic status determines covid-19 incidence and related mortality in santiago, chile. Science, 2021. ISSN 0036-8075. doi: 10.1126/science.abg5298. URL https://science.sciencemag.org/content/early/2021/04/26/science.abg5298.

[9] Ariel Karlinsky and Dmitry Kobak. The world mortality dataset: Tracking excess mortality across countries during the covid-19 pandemic. medRxiv, 2021.

[10] Excess Deaths Associated with COVID-19. https://www.cdc.gov/nchs/nvss/vsrr/covid19/excess_deaths.htm#techNotes, 2020.

[11] CP Farrington, Nick J Andrews, AD Beale, and MA Catchpole. A statistical algorithm for the early detection of outbreaks of infectious disease. Journal of the Royal Statistical Society: Series A (Statistics in Society), 159(3):547–563, 1996.

[12] Angela Noufaily, Doyo G Enki, Paddy Farrington, Paul Garthwaite, Nick Andrews, and Andre Charlett. An improved algorithm for outbreak detection in multiple surveillance systems. Statistics in medicine, 32(7):1206–1222, 2013.

[13] Michael Höhle and Michaela Paul. Count data regression charts for the monitoring of surveillance time series. Computational Statistics & Data Analysis, 52(9):4357–4368, 2008.

[14] Mäelle Salmon, Dirk Schumacher, and Michael Höhle. Monitoring count time series in r: Aberration detection in public health surveillance. 2016.

[15] Jaison R Abel and Richard Deitz. The causes and consequences of puerto rico’s declining population. Current issues in Economics and Finance, 20(4), 2014.

[16] M Smith, K Yourish, S Almukhtar, K Collins, D Ivory, A McCann, et al. Coronavirus in the us: Latest map and case count. The New York Times [Internet].[cited 2020 Apr 1].

[17] Hurricane Maria Updates. In puerto rico, the storm ‘destroyed us’. New York Times. http://www.nytimes.com/2017/09/21/us/hurricane-maria-puerto-rico.html. Accessed February, 15, 2018.

[18] Adam Rogers. In puerto rico, no power means no telecommunications. https://www.wired.com/story/in-puerto-rico-no-power-means-no-telecommunications/, 2017.

[19] John Sandberg, Carlos Santos-Burgoa, Amira Roess, Ann Goldman-Hawes, Cynthia M Pérez, Alejandra Garcia-Meza, and Lynn R Goldman. All over the place?: differences in and consistency of excess mortality estimates in puerto rico after hurricane maria. Epidemiology, 30(4):549–552, 2019.

[20] Nishant Kishore, Domingo Marqués, Ayesha Mahmud, Mathew V Kiang, Irmary Rodriguez, Arlan Fuller, Peggy Ebner, Cecilia Sorensen, Fabio Racy, Jay Lemery, et al. Mortality in puerto rico after hurricane maria. New England journal of medicine, 379 (2):162–170, 2018.

[21] Tyler M Sharp, Kyle R Ryff, Luisa Alvarado, Wun-Ju Shieh, Sherif R Zaki, Harold S Margolis, and Brenda Rivera-Garcia. Surveillance for chikungunya and dengue during the first year of chikungunya virus circulation in puerto rico. The Journal of infectious diseases, 214(suppl 5):S475–S481, 2016.

[22] André Ricardo Ribas Freitas, Maria Rita Donalisio, and Pedro María Alarcón-Elbal. Excess mortality and causes associated with chikungunya, puerto rico, 2014–2015. Emerging infectious diseases, 24(12):2352, 2018.

[23] Shakoor Hajat, Ben G Armstrong, Nelson Gouveia, and Paul Wilkinson. Mortality displacement of heat-related deaths: a comparison of delhi, sao paulo, and london. Epidemiology, pages 613–620, 2005.

[24] Jonathan Dushoff, Joshua B Plotkin, Cecile Viboud, David JD Earn, and Lone Simonsen. Mortality due to influenza in the united states—an annualized regression approach using multiple-cause mortality data. American journal of epidemiology, 163(2):181–187, 2006.

[25] Nazrul Islam, Vladimir M Shkolnikov, Rolando J Acosta, Ilya Klimkin, Ichiro Kawachi, Rafael A Irizarry, Gianfranco Alicandro, Kamlesh Khunti, Tom Yates, Dmitri A Jdanov, et al. Excess deaths associated with covid-19 pandemic in 2020: age and sex disaggre-gated time series analysis in 29 high income countries. bmj, 373, 2021.

[26] Robert William Kates, Craig E Colten, Shirley Laska, and Stephen P Leatherman. Reconstruction of new orleans after hurricane katrina: a research perspective. Proceedings of the national Academy of Sciences, 103(40):14653–14660, 2006.

[27] Martín Echenique and Luis Melgar. Mapping puerto rico’s hurricane migration with mobile phone data. city lab 11 may, 2018.

[28] Jennifer Hinojosa, Nashia Roman, and Edwin Melendez. Puerto rican post-maria relocation by states. Center for Puerto Rican Studies, 1(1):1–15, 2018.

[29] Teralytics. https://www.teralytics.net, 2020.

